# Experiment aversion among clinicians and the public — an obstacle to evidence-based medicine and public health

**DOI:** 10.1101/2023.04.05.23288189

**Authors:** Randi L. Vogt, Patrick R. Heck, Rebecca M. Mestechkin, Pedram Heydari, Christopher F. Chabris, Michelle N. Meyer

## Abstract

**Background:** Randomized controlled trials (RCTs) are essential for determining the safety and efficacy of healthcare interventions. However, both laypeople and clinicians often demonstrate experiment aversion: preferring to implement either of two interventions for everyone rather than comparing them to determine which is best. We studied whether clinician and layperson views of pragmatic RCTs for Covid-19 or other interventions became more positive early in the pandemic, which increased both the urgency and public discussion of RCTs.

**Methods:** We conducted several survey studies with laypeople (total *n*=2,909) and two with clinicians (*n*=895; *n*=1,254) in 2020 and 2021. Participants read vignettes in which a hypothetical decision-maker who sought to improve health could choose to implement intervention A for all, implement intervention B for all, or experimentally compare A and B and implement the superior intervention. Participants rated and ranked the appropriateness of each decision.

**Results:** Compared to our pre-pandemic results, we found no decrease in laypeople’s aversion to non-Covid-19 experiments involving catheterization checklists and hypertension drugs. Nor were either laypeople or clinicians less averse to Covid-19 RCTs (concerning corticosteroid drugs, vaccines, intubation checklists, proning, school reopening, and mask protocols), on average. Across all vignettes and samples, levels of experiment aversion ranged from 28% to 57%, while levels of experiment appreciation (in which the RCT is rated higher than the participant’s highest-rated intervention) ranged from only 6% to 35%.

**Conclusions:** Advancing evidence-based medicine through pragmatic RCTs will require anticipating and addressing experiment aversion among both patients and healthcare professionals.

## Introduction

Randomized controlled trials (RCTs) are crucial for understanding how to safely, effectively, and equitably prevent and treat disease and deliver healthcare. They have repeatedly upended conventional clinical wisdom and the results of observational studies,^1^ and are urgently needed to evaluate new technologies.^2^ However, RCTs often prove controversial, even when they compare interventions that are within the standard of care or otherwise unobjectionable, and about which the relevant expert community is in equipoise.^3^ Prestigious medical journals have recently published several trials—including SUPPORT^4^, FIRST^5^, and iCOMPARE^6^—that have received considerable criticism from physician-scientists, ethicists, and regulators in those journals^7,8^ and the public square.^9–12^ Although criticisms of RCTs can be complex and nuanced, many reflect a rejection of the very idea that an experiment was conducted, as opposed to simply giving everyone the allegedly superior intervention.

In prior studies—inspired by several “notorious RCTs,” including technology industry “A/B tests”^13–15^—we confirmed that substantial shares of both laypeople and clinicians can be averse to randomized evaluation of efforts to improve health. People rated a pragmatic RCT designed to compare the effectiveness of two interventions significantly lower than the average rating of implementing either one, untested, for everyone, a phenomenon we call the “A/B effect.”^16^ In some cases, the lower average rating of an experiment could be driven not by dislike of experiments, per se, but by the fact that many people believe one of its arms is inferior to the other,^16,17^ a belief that is often not evidence-based. We therefore also documented “experiment aversion”: rating an RCT comparing two interventions as worse than *even one’s own least-preferred intervention*.^17^ Both patterns of negative sentiments about experiments—including experiments judged to compare two unobjectionable interventions—can impede efforts to identify what does and does not work to improve health outcomes.

The Covid-19 pandemic presented a potential inflection point in attitudes towards health experimentation. In April 2020, 72 Covid-19 drug trials were already underway^18^ and RCTs became daily, front-page news. That sustained exposure might have educated people about RCTs, or made RCTs more normative. Separately, our previous research suggests that one cause of experiment aversion is an illusion of knowledge—a (mis)perception that experts already must know what works best, and should simply implement that. But Covid-19 was a novel disease, and—at least in the case of pharmaceutical interventions—no sensible person thought the correct treatments were already obvious. People therefore may be less averse to Covid-19 RCTs than to RCTs that test interventions against longstanding conditions or problems. On the other hand, because of the urgency attached to Covid-19, people may be *more* averse to Covid-19 RCTs, being even less inclined to risk giving someone a treatment that might turn out to “lose” in a comparison study.^19,20^ Finally, even if the pandemic did not affect public attitudes towards RCTs, it could have affected the attitudes of clinicians, many of whom were involved in Covid-19 research. Because clinicians strongly influence whether particular RCTs are conducted, their attitudes matter.

We investigated attitudes towards experimentation in the first year of the pandemic by conducting a series of preregistered studies between August 2020 and February 2021. First, we used decision-making vignettes from our previous work to ask whether the extraordinary publicity around Covid-19 RCTs reduced general healthcare experiment aversion by the public. Next, we adapted these vignettes to determine whether the public was averse to experimentation on pharmaceutical and/or non-pharmaceutical interventions (NPIs) for Covid-19. Finally, we recruited two large clinician samples to investigate how their attitudes compared to those of laypeople. All three studies were randomized survey experiments in which participants first read about a decision-maker faced with a problem who either implemented one of two interventions (A or B) or ran an experiment to compare them (and then implemented the superior one). Participants then evaluated how appropriate each of those three decisions was.

## Methods

### Lay Sentiments About Healthcare Experimentation

In August 2020, we used the CloudResearch service to recruit 700 crowd workers on Amazon Mechanical Turk to participate in a brief online survey. These services provide samples that are broadly representative of the U.S. population and are well-accepted in social science research as providing as good or better-quality data than convenience samples such as student volunteers, with results that are similar to probability sampling methods.^21,22^

Each participant first read a vignette that described a problem that the decision-maker could address in three ways (see Table 1 for examples; see the Supplemental Appendix [SA] for text and motivations for all vignettes): by implementing intervention A for all patients (A); by implementing intervention B for all patients (B); or by conducting an experiment in which patients are randomly assigned to A or B and the superior intervention is then implemented for all (A/B). (Our vignettes are silent about whether consent will be obtained, but IRBs customarily waive consent when it would make low-risk pragmatic RCTs impracticable^23^; in separate work, we found that substantial shares of people object to such experiments even when we specify that consent will be obtained.^24^) Next, following standard methods in social and moral psychology for evaluating decisions,^25^ participants rated each option on a scale of appropriateness from 1 (“very inappropriate”) to 5 (“very appropriate”), with 3 as a neutral midpoint. Participants then rank-ordered the options from best to worst. We also collected demographic information, but found no substantial associations with it in any of our studies (Tables S8-11).

**Table 1.** Vignette text for Catheterization Safety Checklist and Ventilator Proning.

|  | (A) Catheterization Safety Checklist | (B) Ventilator Proning |
| --- | --- | --- |
| Background | Some medical treatments require a doctor to insert a plastic tube into a large vein. These treatments can save lives, but they can also lead to deadly infections. | Some coronavirus (Covid-19) patients have to be sedated and placed on a ventilator to help them breathe. Even with a ventilator, these patients can have dangerously low blood oxygenation levels, which can result in death. Current standards suggest that laying ventilated patients on their stomach for 12-16 hours per day can reduce pressure on the lungs and might increase blood oxygen levels and improve survival rates. |
| Intervention A | A hospital director wants to reduce these infections, so he decides to give each doctor who performs this procedure a new ID badge with a list of standard safety precautions for the procedure printed on the back. All patients having this procedure will then be treated by doctors with this list attached to their clothing. | A hospital director wants to save as many ventilated Covid-19 patients as possible, so he decides that all of these patients will be placed on their stomach for 12-13 hours per day. |
| Intervention B | A hospital director wants to reduce these infections, so he decides to hang a poster with a list of standard safety precautions for this procedure in all procedure rooms. All patients having this procedure will then be treated in rooms with this list posted on the wall. | A hospital director wants to save as many ventilated Covid-19 patients as possible, so he decides that all of these patients will be placed on their stomach for 15-16 hours per day. |
| A/B test | A hospital director thinks of two different ways to reduce these infections, so he decides to run an experiment by randomly assigning patients to one of two test conditions. Half of patients will be treated by doctors who have received a new ID badge with a list of standard safety precautions for the procedure printed on the back. The other half will be treated in rooms with a poster listing the same precautions hanging on the wall. After a year, the director will have all patients treated in whichever way turns out to have the highest survival rate. | A hospital director thinks of two different ways to save as many ventilated Covid-19 patients as possible, so he decides to run an experiment by randomly assigning ventilated Covid-19 patients to one of two test conditions. Half of these patients will be placed on their stomach for 12-13 hours per day. The other half of these patients will be placed on their stomach for 15-16 hours per day. After one month, the director will have all ventilated Covid-19 patients treated in whichever way turns out to have the highest survival rate. |

Participants were randomly assigned to one of two vignettes. In Best Anti-Hypertensive Drug, some doctors in a walk-in clinic prescribe “Drug A” while others prescribe “Drug B” (both of which are affordable, tolerable, and FDA approved) and Dr. Jones prescribes either A or B for all his hypertensive patients, or runs a randomized experiment to compare their effectiveness. In Catheterization Safety Checklist, a hospital director similarly considers two locations where he might display a safety checklist for clinicians (see Table 1A).

We define the “A/B Effect” as the degree to which participants’ ratings of the A/B test were lower than the average of their ratings of implementing A and B.^16^ “Experiment aversion” is the degree to which participants rated the A/B test *lower* than *their own lowest-rated* intervention (either A or B for each person). “Experiment appreciation” is the opposite: the degree to which the experiment is rated *higher* than each participant’s *highest-rated* intervention. (See the SA for full details on power analyses and sample sizes, statistical analyses, materials, preregistrations, and data availability.)

### Lay Sentiments About Covid-19 Healthcare Experimentation

Between August 2020 and January 2021, we recruited 2,209 additional laypeople in the same manner described above. They read, rated, and ranked six new vignettes involving Covid-19 interventions (N = 339–450 per vignette). Four vignettes were based on Covid-19-related interventions that were discussed, tested, and/or implemented at the time: Masking Rules (which described two masking policies, of varying scope); School Reopening (two school schedules designed to increase social distancing); Best Vaccine (two types of vaccine—mRNA versus inactivated virus); and Ventilator Proning (two protocols for positioning ventilated Covid-19 patients; see Table 1B). The other two vignettes—Intubation Safety Checklist and Best Corticosteroid Drug—were adapted from the first study to apply to Covid-19.

### Clinician Sentiments About Covid-19 Healthcare Experimentation

Between November 2020 and February 2021, clinicians (including physicians, physician assistants, and nurse practitioners) in a large health system in the Northeastern U.S. were recruited by email to read, rate, and rank one of four Covid-19-related vignettes (Masking Rules: *n* = 349; Intubation Safety Checklist: *n* = 271; Best Corticosteroid Drug: *n* = 275; Best Vaccine: *n* = 1254) from the second study.

## Results

### Lay Sentiments About Healthcare Experimentation

We found substantial negative reactions to A/B testing in both vignettes (Table 2A), replicating our pre-pandemic findings.^16,17^ In Catheterization Safety Checklist (Figure 1A), we found evidence of the A/B Effect: participants rated the A/B test significantly below the average ratings they gave to implementing interventions A and B (*d* = 0.69, 95% CI: (0.53, 0.85)). Here, 41% ± 5% (95% CI) of participants expressed experiment aversion (rating the A/B test lower than their own lowest-rated intervention; *d* = 0.25, 95% CI: (0.11, 0.39)). When ranking the three options from best to worst, only 32% placed the A/B test first, while 48% placed it last.

**Figure 1.**
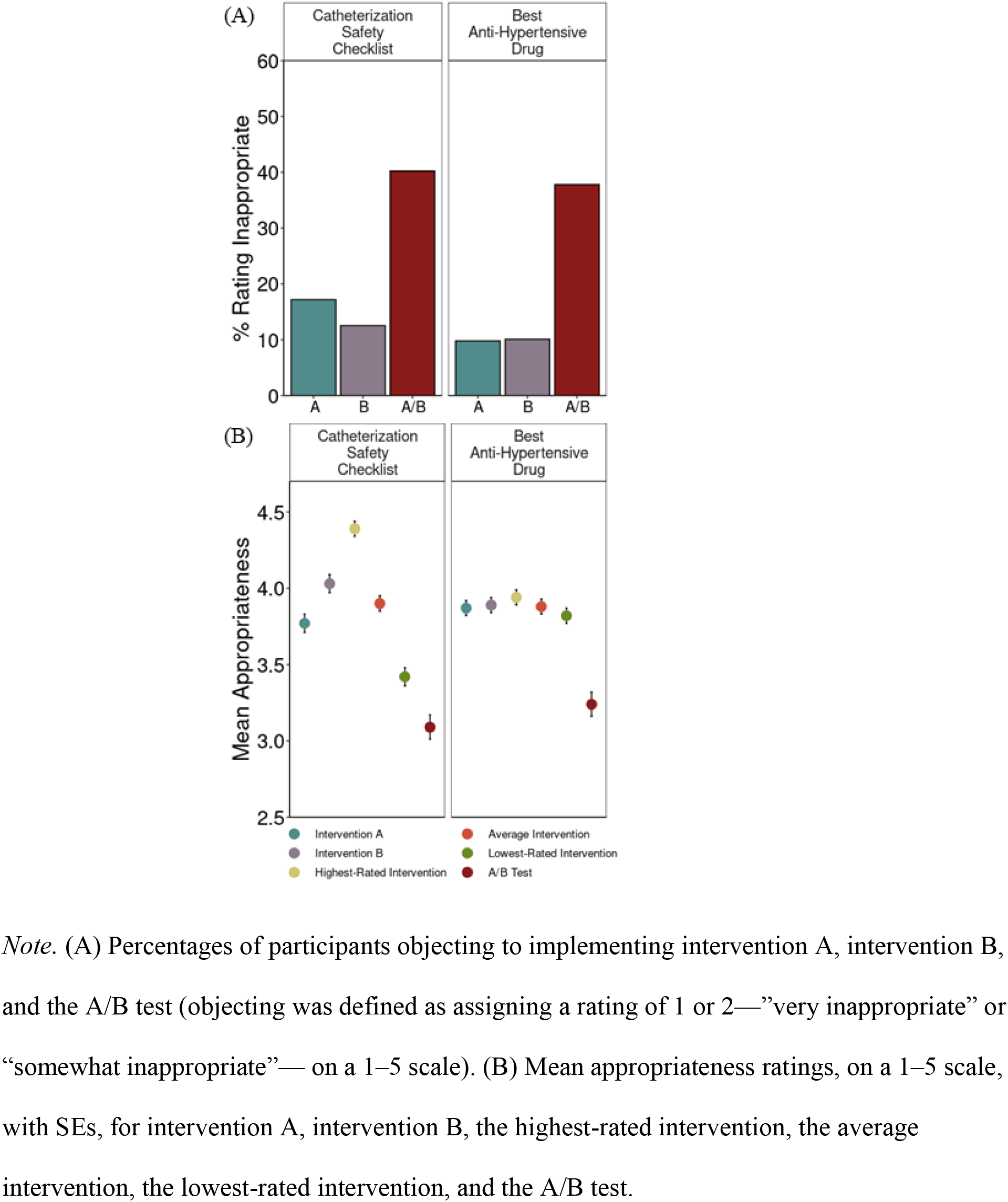
Lay Sentiments About Healthcare Experimentation.

**Table 2.** Sentiments about experiments by vignette and population.

|  | Negative sentiment |  |  |  | Positive sentiment |  |  |  |
| --- | --- | --- | --- | --- | --- | --- | --- | --- |
|  | Experiment Aversion | A/B Effect | More people averse than appreciative? | More people rank AB test worst than best? | More people rank AB test best than worst? | More people appreciative than averse? | Reverse A/B Effect | Experiment Appreciation |
| **(A) Lay Sentiments About Healthcare Experimentation** |  |  |  |  |  |  |  |  |
| Catheterization Safety Checklist | ✓ | ✓ | ✓ | ✓ |  |  |  |  |
| Best Anti-Hypertensive Drug | ✓ | ✓ | ✓ |  |  |  |  |  |
| **(B) Lay Sentiments About Covid-19 Healthcare Experimentation** |  |  |  |  |  |  |  |  |
| Ventilator Proning | ✓ | ✓ | ✓ |  |  |  |  |  |
| School Reopening |  | ✓ | ✓ | ✓ |  |  |  |  |
| Masking Rules | ✓ | ✓ | ✓ | ✓ |  |  |  |  |
| Intubation Safety Checklist | ✓ | ✓ | ✓ | ✓ |  |  |  |  |
| Best Corticosteroid Drug |  | ✓ |  |  | ✓ |  |  |  |
| Best Vaccine |  | ✓ |  |  | ✓ |  |  |  |
| **(C) Clinician Sentiments About Covid-19 Healthcare Experimentation** |  |  |  |  |  |  |  |  |
| Masking Rules | ✓ | ✓ | ✓ | ✓ |  |  |  |  |
| Intubation Safety Checklist | ✓ | ✓ | ✓ | ✓ |  |  |  |  |
| Best Corticosteroid Drug | ✓ | ✓ | ✓ |  |  |  |  |  |
| Best Vaccine |  | ✓* |  |  | ✓ |  |  |  |

*Note.* The A/B Effect refers to the difference between the average rating of the two interventions and the rating of the A/B test. Experiment Aversion refers to the difference between the lowest-rated intervention and the rating of the A/B test. The Reverse A/B Effect refers to the difference between the rating of the A/B test and the average rating of the two interventions. Experiment Appreciation refers to the difference between the rating of the A/B test and the rating of the highest-rated intervention.
Checkmarks (✓) represent a statistically significant effect at $p < .05$ . In one case, the checkmark is followed by an asterisk (\*). This indicates that while the effect reaches statistical significance, the effect size is very small and might have only reached significance due to the large sample size (three times as large as that for other vignettes).
Variables to the right of the thick vertical line are the reverse of those on the left. If no checkmark appears in either of the corresponding columns to the left and right of the thick vertical line (e.g., "More people rank A/B test worst than best?" and "More people rank A/B test best than worst?"), that means that there is no significant difference (e.g., there is no statistically significant difference between the proportion of people ranked that A/B test worst and the proportion of people who ranked the A/B test best).

We also observed an A/B Effect in Best Anti-Hypertensive Drug (Figure 1B); *d* = 0.52, 95% CI: (0.36, 0.68)), where 44% ± 5% also expressed experiment aversion (*d* = 0.46, 95% CI: (0.30, 0.52)). Notably, participants were averse to this experiment even though there is no reason to prefer “Drug A” to “Drug B,” and patients are effectively already randomized to A or B based on which clinician happens to see them—which occurs wherever unwarranted variation in practice determines treatments, such as walk-in clinics and emergency departments. Here, however, similar proportions of people ranked the A/B test best and worst (50% vs. 45%; *p* = 0.16).

These levels of experiment aversion near the height of the pandemic were slightly (but not significantly) higher than those we observed among similar laypeople in 2019 (41% ± 5% in 2020 vs. 37% ± 6% in 2019 for Catheterization Safety Checklist, *p* = 0.31 ; 44% ± 5% in 2020 vs. 40% ± 6% in 2019 for Best Anti-Hypertensive Drug, *p* = 0.32).^17^

### Lay Sentiments About Covid-19 Specific Healthcare Experimentation

In all six Covid-19 vignettes, we found evidence of the A/B Effect (Table 2B). In three, however, we did not find experiment aversion: Best Vaccine, Best Corticosteroid Drug, and School Reopening. In the first two of these, participants rated the two interventions very similarly and the experiment only slightly lower (Figure 2B). These vignettes also elicited the largest proportion of participants (65% in Best Vaccine and 56% in Best Corticosteroid Drug) in any vignette who ranked the A/B test best among the three options, compared to 31–34% of participants who ranked it worst. In School Reopening, experiment aversion was not observed because participants on average clearly preferred intervention B to A and rated the experiment similar to intervention A.^26,27^ 53% of participants ranked intervention B as the best of the three options (compared to 17% choosing intervention A and 30% choosing the A/B test).

**Figure 2.**
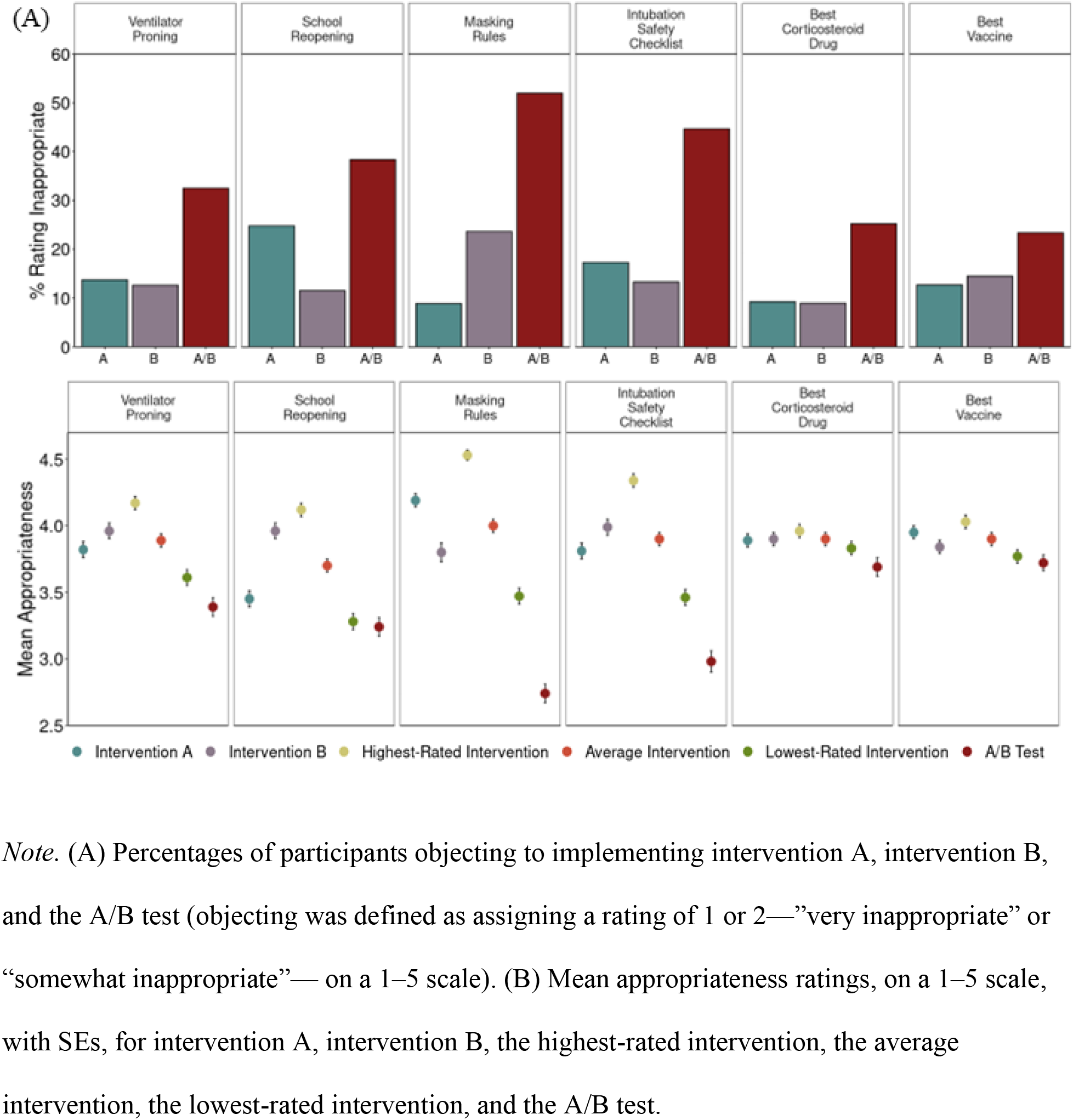
Lay Sentiments About Covid-19 Specific Healthcare Experimentation.

In the other three vignettes, participants rated the A/B test condition as significantly less appropriate than their lowest-rated intervention (Masking Rules: *d* = 0.56, 95% CI: (0.41, 0.71); Ventilator Proning: *d* = 0.17, 95% CI: (0.04, 0.30); Intubation Safety Checklist: *d* = 0.36, 95% CI: (0.21, 0.49)). These levels of aversion to Covid-19 RCTs are similar to the levels of aversion to non-Covid-19 RCTs both before^17^ and during the pandemic (see above).

### Clinician Sentiments About Covid-19 Specific Healthcare Experimentation

We observed an A/B effect in all four vignettes. In two, clinicians, like laypeople, were also significantly experiment averse (Masking Rules: *d* = 0.74, 95% CI: (0.57, 0.91); Intubation Safety Checklist: *d* = 0.30, 95% CI: (0.15, 0.45)). In Best Vaccine, clinicians, like laypeople, did not show any significant difference in their ratings of the A/B test and their lowest-rated intervention (*d* = –0.03, 95% CI: (–0.10, 0.04)). Again, like laypeople, 58% of clinicians ranked the vaccine A/B test as the best of the three options, the highest proportion of any clinician-rated vignette.

Clinicians differed from laypeople in their response to Best Corticosteroid Drug. Laypeople did not show experiment aversion, but clinicians rated the A/B test as significantly less appropriate than their lowest-rated intervention (*d* = 0.49, 95% CI: (0.32, 0.66)). This difference may be due to clinicians’ greater familiarity with the treatment of Covid-19. Clinicians may also have seen an urgent need for *any* drugs to treat Covid-19^20^ and thus rated adopting a clear treatment intervention as more appropriate than an RCT.

**Figure 3.**
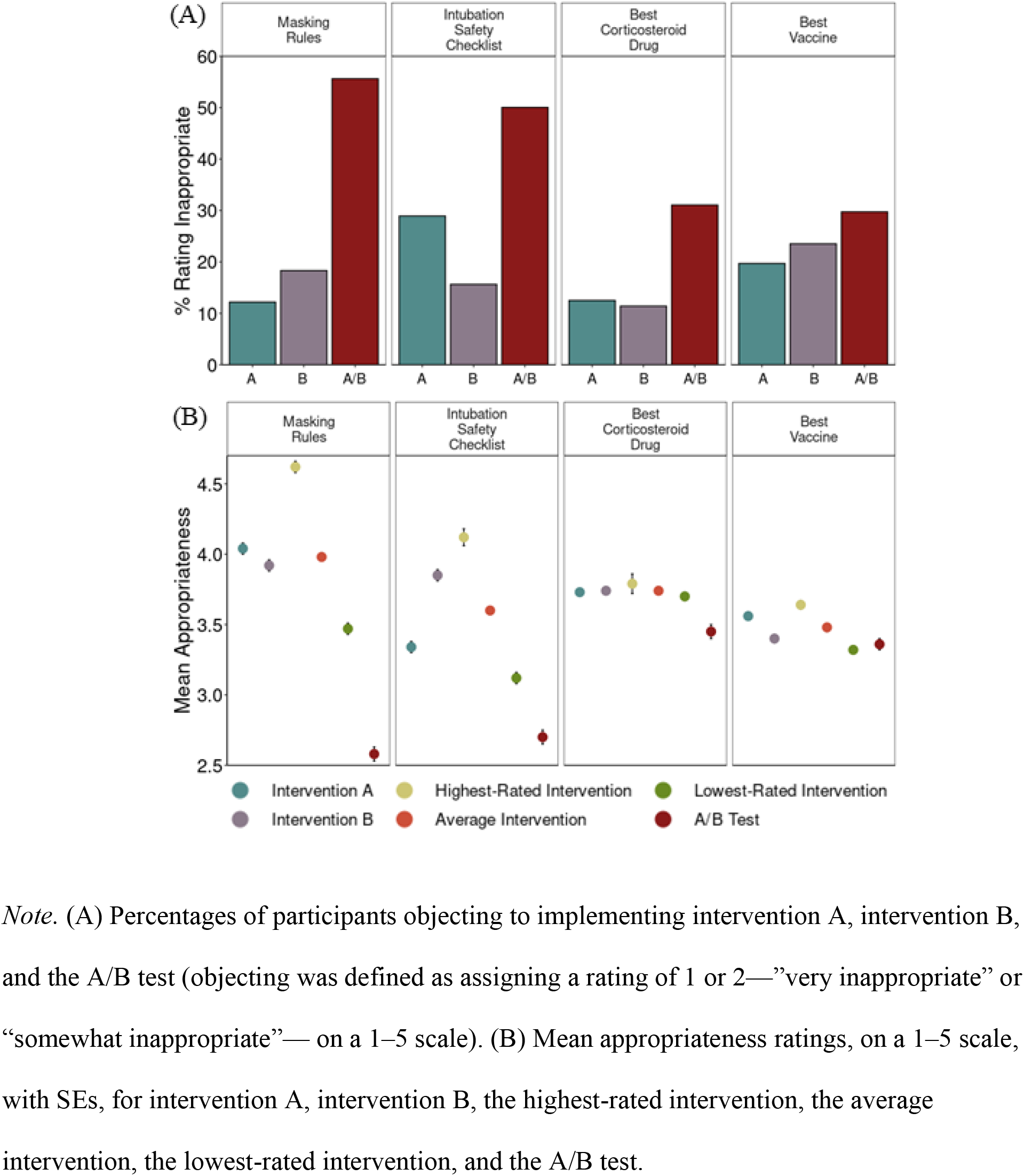
Clinician Sentiments About Covid-19 Specific Healthcare Experimentation.

## Discussion

We found no diminution in general experiment aversion among laypeople during the first year of the Covid-19 pandemic, despite increased exposure to the nature and purpose of RCTs. Neither laypeople nor clinicians were overall less averse to Covid-19 experiments, despite the fact that confidence in anyone’s knowledge of what works should have been even more circumscribed than in the everyday contexts of hypertension and catheter infections. To the contrary, we found an A/B effect (the average rating of the RCT was lower than the average rating of the two policies) in all vignettes and samples. Most Covid-19 vignettes were met with experiment aversion (on average, participants rated the RCT lower than each participant’s lowest-rated intervention). This is consistent with an emphasis during the pandemic that we must “do” instead of “learn,” a false dichotomy that fails to recognize that implementing an untested intervention is itself a nonconsensual experiment from which, unlike an RCT, little or nothing can be learned.^28–30^ Similarly, across *all* vignettes and samples, between 28% and 57% of participants demonstrated experiment aversion, while only 6%–35% demonstrated experiment appreciation (by rating the RCT higher than their highest-rated intervention).

In none of our 12 studies were more people appreciative of than averse to the RCT, in none was the average RCT rating higher than the average intervention rating, and in none was the RCT rating higher than each participant’s highest-rated intervention, on average. Notably, unlike trials with placebo or no-contact controls, the A/B tests in our vignettes compared two active, plausible interventions, neither of which was obviously known ex ante to be superior. Yet substantial shares of participants still preferred that one intervention simply be implemented without bothering to determine which (if either) worked best.

There is one bright spot in our results: the most positive sentiment towards experiments was observed in both laypeople and clinicians in the vignettes involving Covid-19 drugs and vaccines. Here we observed the highest proportions of participants who demonstrated experiment appreciation (31%–46%) and who ranked the RCT first (49%–65%). This result is consistent with our previous findings that the illusion of knowledge—here, the belief that either the participant herself or some expert already does or should know the right thing to do and should simply do it—biases people to prefer universal intervention implementation to RCTs.^16,17^ Rightly or wrongly, both laypeople and clinicians might (a) appropriately recognize that near the start of a pandemic, no one knows which existing drugs, if any, are safe and effective in treating a novel disease, and that new vaccines need to be tested, yet (b) fail to sufficiently appreciate the level of uncertainty around NPIs like masking, proning, and social distancing, which can also benefit from rigorous evaluation. This is consistent with the dearth of RCTs of Covid-19 NPIs:^31^ of the more than 4,000 Covid-19 trials registered worldwide as of August 2021, only 41 tested NPIs.^32^ Explaining critical concepts like clinical equipoise or unwarranted variation in medical and NPI practice alike might diminish experiment aversion.

Critics note that RCTs have limited external validity when they employ overly selective inclusion/exclusion criteria or are executed in ways that deviate from how interventions would be operationalized in diverse, real-world settings. However, the solution is not to abandon randomized evaluation, but to incorporate it into routine clinical care and healthcare delivery via pragmatic RCTs.^1,33^ It has been many years since the Institute of Medicine urged research of many varieties to be embedded in care.^34^ More recently, the FDA established a Real-World Evidence Program that promotes pragmatic RCTs to support post-marketing monitoring and other regulatory decision-making.^35,36^ Pragmatic RCTs have been fielded successfully and informed healthcare practice and policy,^37–39^ but they remain far from ubiquitous and they require buy-in to be successful, as shown by the case of a Denmark school reopening trial that was abandoned due to lack of such support.^40^ Wider use of pragmatic RCTs will require not only redoubling investment in interoperable electronic health records and recalibrating regulators’ views of the comparative risks of research versus idiosyncratic practice variation,^1^ but also anticipating and addressing experiment aversion among patients and healthcare professionals.

## Data Availability

Participant response data, preregistrations, materials, and analysis code have been deposited in Open Science Framework and will be released upon final publication of this paper.

## Acknowledgements

Supported by Office of the Director, National Institutes of Health (NIH) (3P30AG034532-13S1) and funded by the Food and Drug Administration (FDA). The content is solely the responsibility of the authors and does not necessarily represent the official views of the NIH or the FDA. We thank Daniel Rosica and Tamara Gjorgjieva for excellent research assistance. Vogt and Heck contributed equally to this work. Meyer and Chabris contributed equally to this work.

## Methods

In the main text, we grouped the vignettes thematically into three sets: “Lay Sentiments About Healthcare Experimentation,” “Lay Sentiments About Covid-19 Specific Healthcare Experimentation,” and “Clinician Sentiments About Covid-19 Specific Healthcare Experimentation.” However, when we collected data, we grouped our vignettes differently such that we started with vignettes that we have used in previous published work and their respective Covid-19 derivatives, then we developed and tested novel Covid-19 specific vignettes separately, and then, again separately, we tested a Covid-19 vaccine vignette. We followed a similar pattern in our clinician sample: we first tested three Covid-19 specific vignettes (two which were derivatives of vignettes from our previous work, one which was new to this work) and then separately, we tested a Covid-19 vaccine vignette. These groupings are important for understanding how participants were randomly assigned to vignettes and why there are slight discrepancies (or large discrepancies in the case of the Best Vaccine vignette in the clinician sample^1^) in the number of participants in each vignette (see Table S1).

**Table S1.**
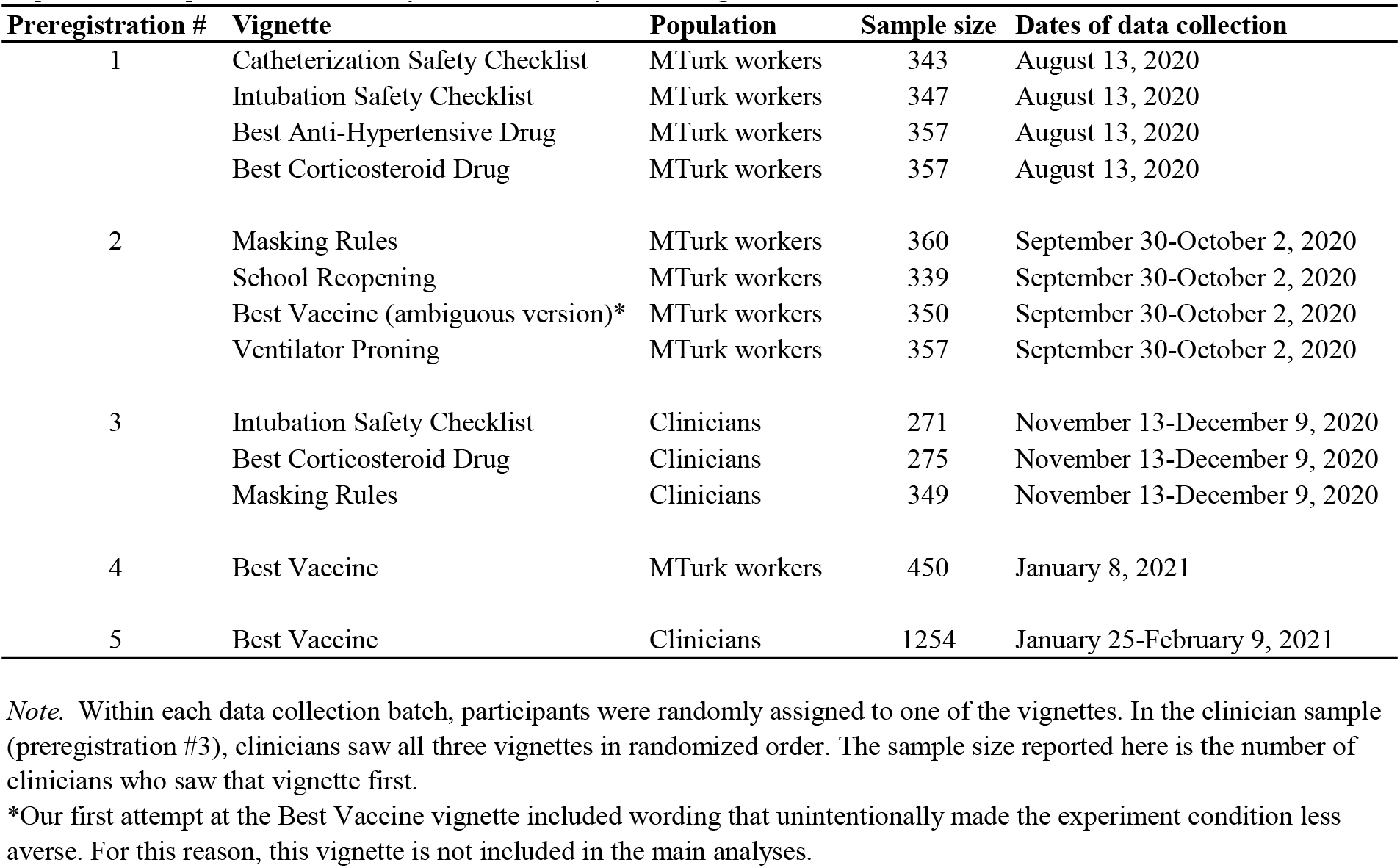
Population, sample size, and dates of data collection for each vignette.

| Preregistration # | Vignette | Population | Sample size | Dates of data collection |
| --- | --- | --- | --- | --- |
| 1 | Catheterization Safety Checklist | MTurk workers | 343 | August 13, 2020 |
|  | Intubation Safety Checklist | MTurk workers | 347 | August 13, 2020 |
|  | Best Anti-Hypertensive Drug | MTurk workers | 357 | August 13, 2020 |
|  | Best Corticosteroid Drug | MTurk workers | 357 | August 13, 2020 |
| 2 | Masking Rules | MTurk workers | 360 | September 30-October 2, 2020 |
|  | School Reopening | MTurk workers | 339 | September 30-October 2, 2020 |
|  | Best Vaccine (ambiguous version)* | MTurk workers | 350 | September 30-October 2, 2020 |
|  | Ventilator Proning | MTurk workers | 357 | September 30-October 2, 2020 |
| 3 | Intubation Safety Checklist | Clinicians | 271 | November 13-December 9, 2020 |
|  | Best Corticosteroid Drug | Clinicians | 275 | November 13-December 9, 2020 |
|  | Masking Rules | Clinicians | 349 | November 13-December 9, 2020 |
| 4 | Best Vaccine | MTurk workers | 450 | January 8, 2021 |
| 5 | Best Vaccine | Clinicians | 1254 | January 25-February 9, 2021 |

*Note.* Within each data collection batch, participants were randomly assigned to one of the vignettes. In the clinician sample (preregistration #3), clinicians saw all three vignettes in randomized order. The sample size reported here is the number of clinicians who saw that vignette first.
\*Our first attempt at the Best Vaccine vignette included wording that unintentionally made the experiment condition less aversive. For this reason, this vignette is not included in the main analyses.
<sup>1</sup> The Best Vaccine vignette was combined with another study that required a sample size much larger than the sample sizes in our previous vignette studies to have adequate statistical power.

For clarity, in the main text of this article we used different names for the vignettes than those used in the preregistrations and in previous publications (see Table S2).

**Table S2.**
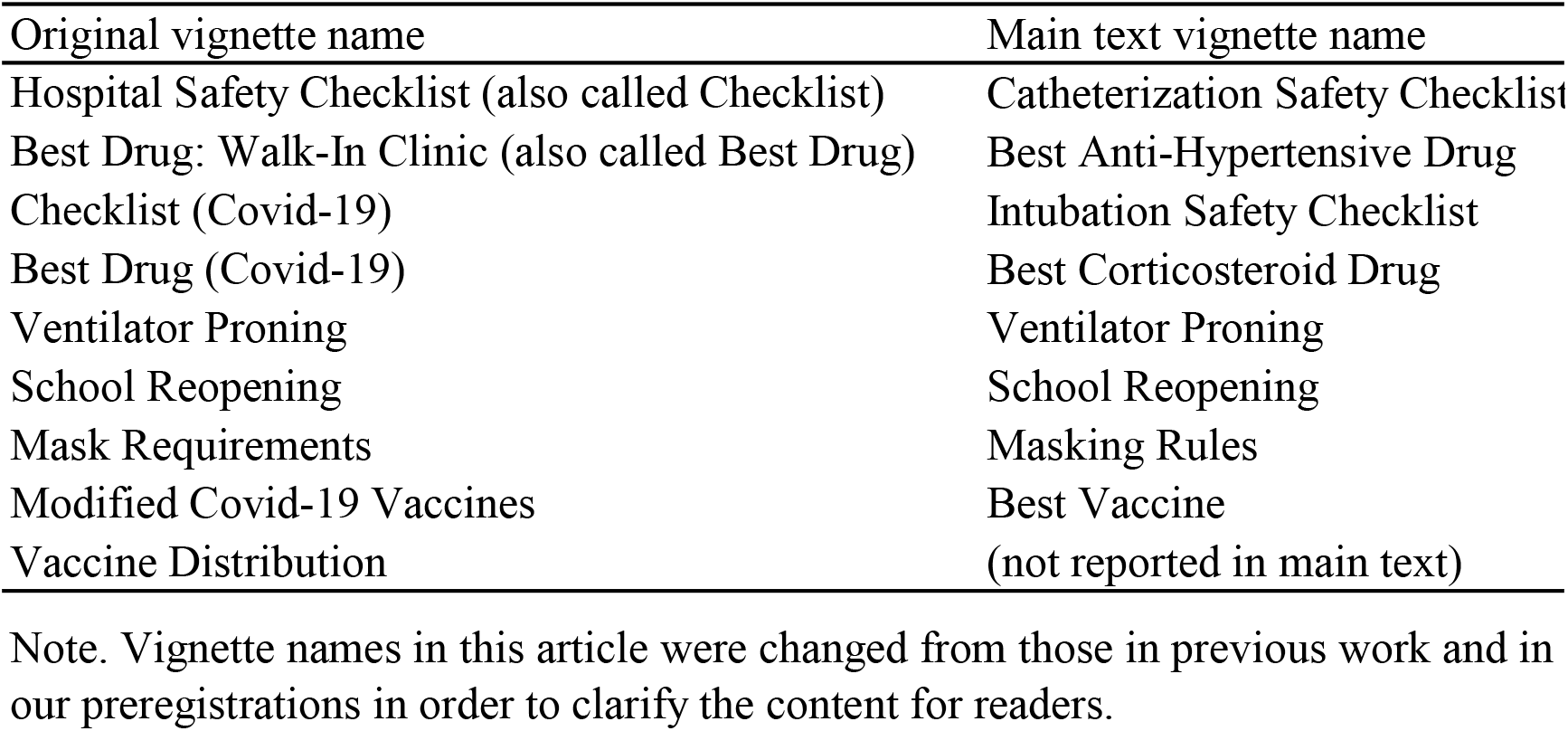
Original vignette names from preregistrations and previous work and corresponding name in main text.

### Preregistrations, sample sizes, and power analyses

Our research questions, power analyses and sample sizes, and analysis plans were all preregistered at Open Science Framework (OSF) before data collection. These sample size precommitments are copied from each preregistration document which will be released upon final publication of this paper.

Preregistration 1 (Catheterization Safety Checklist, Best Anti-Hypertensive Drug, Intubation Safety Checklist, Best Corticosteroid Drug vignettes):

“We predict that, using a two-tailed, paired t-test with ⍺ = .05 within each scenario, participants will rate the A/B test condition as significantly less appropriate than their own average rating of the two policy conditions, mean(A,B). This is the test for the “A/B Effect.” Recruiting 350 participants for each scenario provides 95% power to detect an effect as small as d = 0.19, which is substantially smaller than the effect sizes we have observed using the Hospital Safety Checklist and Best Drug: Walk-In Clinic vignettes in past research.”

Preregistration 2 (Ventilator Proning, School Reopening, Masking Rules, and Best Vaccine (initial ambiguous version) vignettes):

“We predict that, using a two-tailed, paired t-test with ⍺ = .05 within each scenario, participants will rate the A/B test condition as significantly less appropriate than their own average rating of the two policy conditions, mean(A,B). This is the test for the “A/B Effect.” Recruiting 350 participants for each scenario provides 95% power to detect an effect as small as d = 0.19, which is substantially smaller than the effect sizes we have observed using the Hospital Safety Checklist and Best Drug: Walk-In Clinic vignettes in past research.”

Preregistration 3 (Clinicians; Intubation Safety Checklist, Best Corticosteroid Drug, and Masking Rules vignettes):

Note that because of time constraints around the possible starting dates of our clinician surveys, we launched this study before preregistering it, and we did not report an explicit power analysis before collecting the data. Because this study follows a similar structure to the studies above, however, it was reasonable to apply the previous sample size and power analysis considerations. We did, however, preregister our approach and research plan twice during this study: once during data collection, before any analyses had been conducted, and again after all data had been collected (but before analyzing any of them).

> Preregistration 3.1: “At the time of this preregistration, we have received 655 complete responses. No data have been explored or analyzed at this point. We will conduct an interim analysis on this dataset using the same analyses we have previously preregistered, and we may continue to collect more data from this population.”
>
> Preregistration 3.2: “Data collection is now complete and we have closed the survey. On 11/24/2020, we conducted an interim analysis on 601 complete responses. Since then, we have received an additional 295 complete responses, to which we remain blind.”

Preregistration 4 (Best Vaccine):

“We recruited 350 participants for the original Covid-19 vaccines study. Because we are running this study to determine whether even a small effect emerges, we will increase the sample size to 450 participants. This provides 80% power to detect an effect as small as d = 0.13 in a repeated-measures, two-tailed t-test, and 95% power to detect an effect as small as d = 0.17.”

Preregistration 5 (Clinicians; Best Vaccine):

“Our previous survey of healthcare providers resulted in approximately 900 complete responses; we expect a similar response rate for this survey. This sample size provides 95% power to detect an effect as small as d = 0.12 using a two-tailed, repeated measures t-test. Even if we only receive 600 complete responses, we will have 95% power to detect an effect as small as d = 0.15.”

### Procedure and design

Several aspects of the procedure and experimental design were consistent across the studies reported here. Below, we describe these consistent features and note in specific studies where we deviated from them.

For the lay participant samples, we used the CloudResearch service to recruit crowd workers on Amazon Mechanical Turk (MTurk) to participate in a 3–5-minute survey experiment.

Participants were excluded from recruitment in any of the studies reported here if they had participated in any of our previous studies on this topic. Across all laypeople vignettes, the completion rate of participants starting the survey was 91.5%. The Geisinger IRB determined that these anonymous surveys were exempt (IRB# 2017-0449).

For the clinician samples, we recruited healthcare providers from a large health system in the Northeastern U.S via email. Each provider received either one or two emails about the study during the recruitment window. In the first clinician study (Intubation Safety Checklist, Best Corticosteroid Drug, and Masking Rules vignettes), we first tested the email recruitment system by sending out the survey invitation email to just 200 clinicians. Clinicians who completed the survey based on this survey invitation were included in the final sample. Then, all clinicians were sent the recruitment email on November 19, 2020, followed by a reminder email on December 3, 2020. In the second clinician study (Best Vaccine), the initial recruitment email was sent January 25, 2021, with the follow-up email sent February 2, 2021. In the first clinician study, 5,925 clinicians were emailed and 895 completed the survey. In the second clinician study, 6,993 clinicians were emailed and 1,254 completed the survey. In these samples, because survey responses were fully anonymous, we were not able to restrict participation based on our previous studies, so some participants who completed the Best Vaccine vignette may have earlier completed the Intubation Safety Checklist, Best Corticosteroid Drug, and Masking Rules vignettes.

In all cases, participants completed an online survey hosted by Qualtrics. After opening the survey, participants were randomly assigned to one of the possible vignettes being studied.^2,3^ In the case of data collection batches 4 and 5, there was only one vignette being tested that all participants saw. At this point, we used the exact same procedure detailed in Heck et al. (2020)^1^. First, participants were instructed to read about several possible decisions made by different decision-makers^4^, and to try to treat each decision as separate from the others. All scenarios contained a brief “background” text at the top of the page that summarized a problem, followed by three “situations,” each of which detailed the decision-maker’s choice to adopt intervention A, intervention B, or to run an A/B test by randomly assigning people to one of two test conditions. These conditions were presented in fully counterbalanced order; each participant received one of six possible orders (i.e., Situation 1 = A, Situation 2 = B, and Situation 3 = A/B; Situation 1 = A/B, Situation 2 = B, and Situation 3 = A; etc.…). At no point did we observe a meaningful effect of presentation order, so we collapsed across this variable for all analyses.

For our primary outcome measures, participants were asked to rate the appropriateness of the decisions made in Situation 1, Situation 2, and Situation 3 (“How appropriate is the director’s decision in Situation 1/2/3?”), using a 1-5 scale (1 = “Very inappropriate”, 2 = “Inappropriate”, 3 = “Neither inappropriate nor appropriate”, 4 =”Appropriate”, 5 = “Very appropriate”). Participants then specified a ranked order of the three decisions (“Among these three decisions, which decision do you think the director should make? Please drag and drop the options below into your preferred order from best to worst. You must click on at least one option before you can proceed.”), with 1 being the best decision and 3 being the worst. The last item on this page asked participants to explain why they chose these ratings and rankings in a couple of sentences (“In a couple of sentences, please tell us why you chose the ratings and rankings you chose.”).

Following these primary measures, participants completed standard demographic items on the next page. For MTurk participants, these were measures of sex, race/ethnicity, age, educational attainment, household income, religious belief or affiliation, whether they have a degree in a STEM field or not, and four items identifying political orientation and affiliation. As part of an ongoing study in our laboratory (whose results will be reported elsewhere), these participants were randomized to one of six conditions for this demographic questionnaire where we varied the option to select “prefer not to answer” and whether the items were mandatory, optional, or requested (but not required). For clinician participants, demographic items were mandatory response and were limited to the following: sex, sources of training in research methods and statistics, self-reported comfort with research methods and statistics, past experience with activities related to research methods and statistics (e.g., publishing a scientific paper or analyzing data), current involvement in research, position (e.g., doctor, physician assistant, nurse, medical student, etc.), length of time working in the medical field, and field of specialty.

After completing the survey, MTurk participants were given a completion code to receive payment ($0.40). Clinician participants were invited to enter into a lottery to win a $50 Amazon gift card by following a link to an independent survey where they could enter their email address. All participants were thanked for their participation and offered the opportunity to comment on the survey.

### Measures

We computed several variables to measure participants’ sentiments about experimentation.

Following Meyer et al. (2019)^1^, we define an “A/B effect” as the difference between participants’ mean policy rating and their rating of the A/B test—that is, the degree to which the policies are (on average) rated higher than the A/B test. We also report the percentage of participants whose mean policy rating is higher than their rating of the A/B test.

Following Heck et al. (2020^2^; see also Mislavsky et al., 2019^3^), we define “experiment aversion” as the difference between participants’ rating of their own lowest-rated policy and their rating of the A/B test. We also report the percentage of participants who express experiment aversion.

“Experiment rejection” (first reported in Heck et al., 2020^2^, but without this name) occurs when a participant rates the A/B test as inappropriate (1 or 2 on the 5-point scale) while also rating each policy as neutral or appropriate (3–5 on the scale).

A “reverse A/B effect” is the difference between participants’ rating of the A/B test and their mean policy rating—that is, the degree to which the A/B test is rated higher than the policies (on average). We also report the percentage of participants whose rating of the A/B test is higher than their mean policy rating.

“Experiment appreciation” is the difference between participants’ rating of the A/B test and their rating of their own highest-rated policy. We also report the percentage of participants who express experiment appreciation.

“Experiment endorsement” occurs when a participant rates the A/B as appropriate (4 or 5 on the 5-point scale) while also rating each intervention as neutral or inappropriate (1–3 on the scale).

In all cases where a *d-*value was calculated (i.e., A/B effect, experiment aversion, reverse A/B effect, experiment appreciation), we used Cohen’s *d* recovered from the *t*-statistic, *n*, and correlation between the two measures being compared (Dunlop et al., 1996, equation 3^4^: d = *t*_c_[2(1-*r*)/*n*]^½^; see also http://jakewestfall.org/blog/index.php/category/effect-size/kewestfall.org^5^. To calculate this *d*-value, we use the following R code: effsize::cohen.d(x,y, paired = TRUE).

### Vignettes

Our vignettes were inspired by discussions about the ethics of real-world RCTs (see Table S3).

**Table S3.** Literature calling for or reporting an RCT similar to what is proposed in each vignette.

| Vignette name | Relevant literature |
| --- | --- |
| Catheterization Safety Checklist | Pronovost et al.,<sup>6</sup> Urbach et al.,<sup>7</sup> Arriaga et al.<sup>8</sup> |
| Best Anti-Hypertensive Drug | ROMP Ethics Study,<sup>9</sup> Sinnott et al.<sup>10</sup> |
| Intubation Safety Checklist | Turner et al.<sup>11</sup> |
| Best Corticosteroid Drug | Wagner et al.<sup>12</sup> |
| Ventilator Proning | Elharrar et al.,<sup>13</sup> Sartini et al.,<sup>14</sup> Caputo et al.<sup>15</sup> |
| School Reopening | Fretheim et al.<sup>16,17</sup>, Helsingen et al.<sup>18</sup>, Angrist et al.<sup>19</sup>, Kolata<sup>20</sup> |
| Masking Rules | Abaluck et al.<sup>21</sup>, Jefferson et al.<sup>22</sup>, Bundgaard et al.<sup>23</sup> |
| Best Vaccine | Bach<sup>24</sup> |

## Results

### Sample demographics

#### Lay participants

Across all vignettes reported in the main text (i.e., excluding the initial ambiguous version of the Best Vaccine vignette), there were a total of 2,910 lay participants. They ranged in age from 18 to 88 years old (mean = 38.4, SD = 12.8) and the majority were White (74.6%) and female (55.9%). 35.7% had a 4-year college degree, 29.7% had some college, and 20.5% had a graduate degree. 21.3% of participants had a degree in a STEM field. The most frequently selected income level was between $20,000 and $40,000 (20.7%). A majority of participants reported being moderate, leaning liberal, or being liberal both generally and specifically with regards to social and economic issues. Similarly, a majority of participants reported being independent, leaning Democrat, or being Democrat in their political party affiliations. 37.7% of participants reported being non-religious. Of those who reported being religious, the most reported religion was Protestant (24.2%). See Table S4 for demographic breakdowns by vignette and in the combined lay participant sample.

**Table S4.**
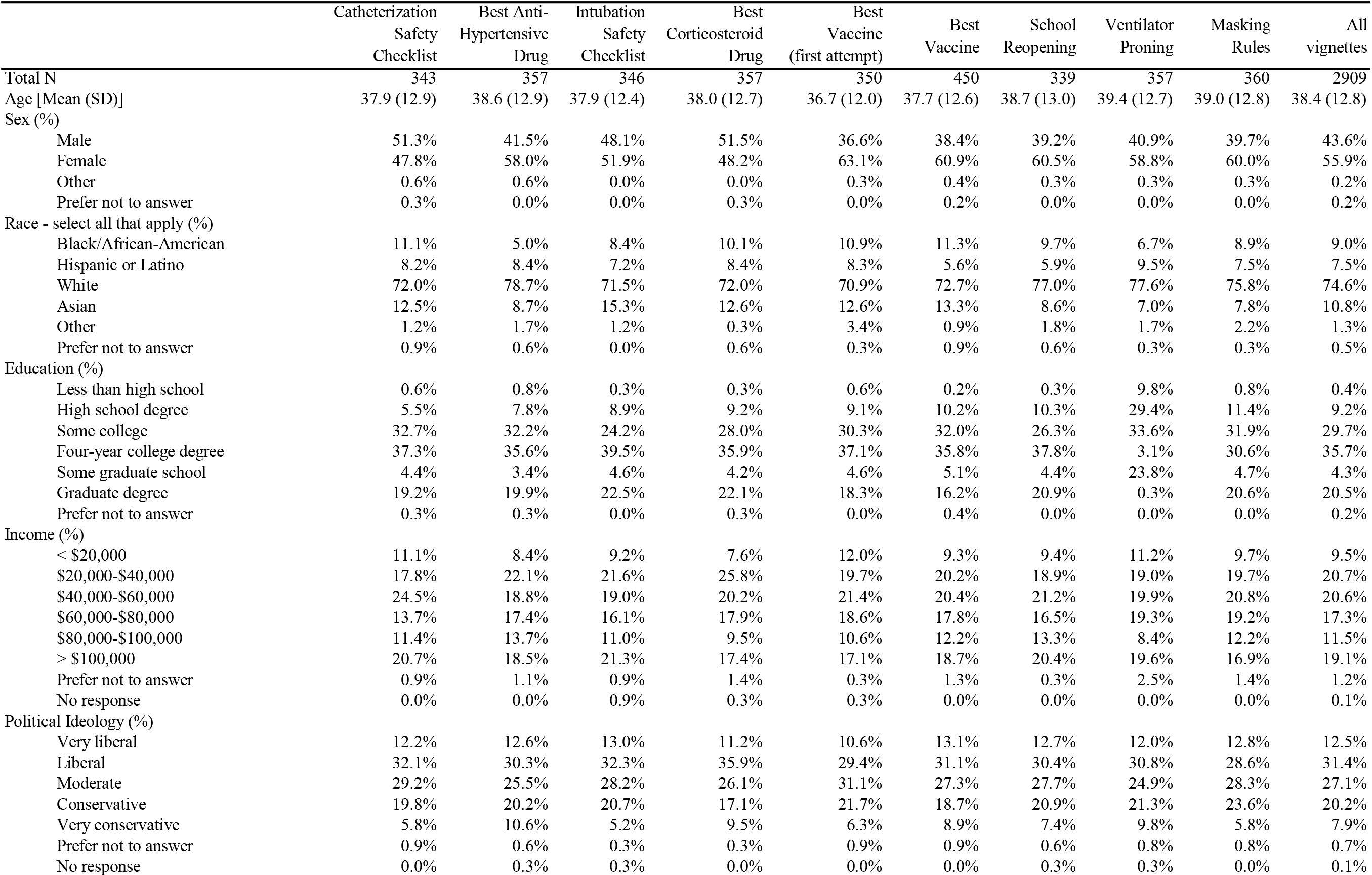
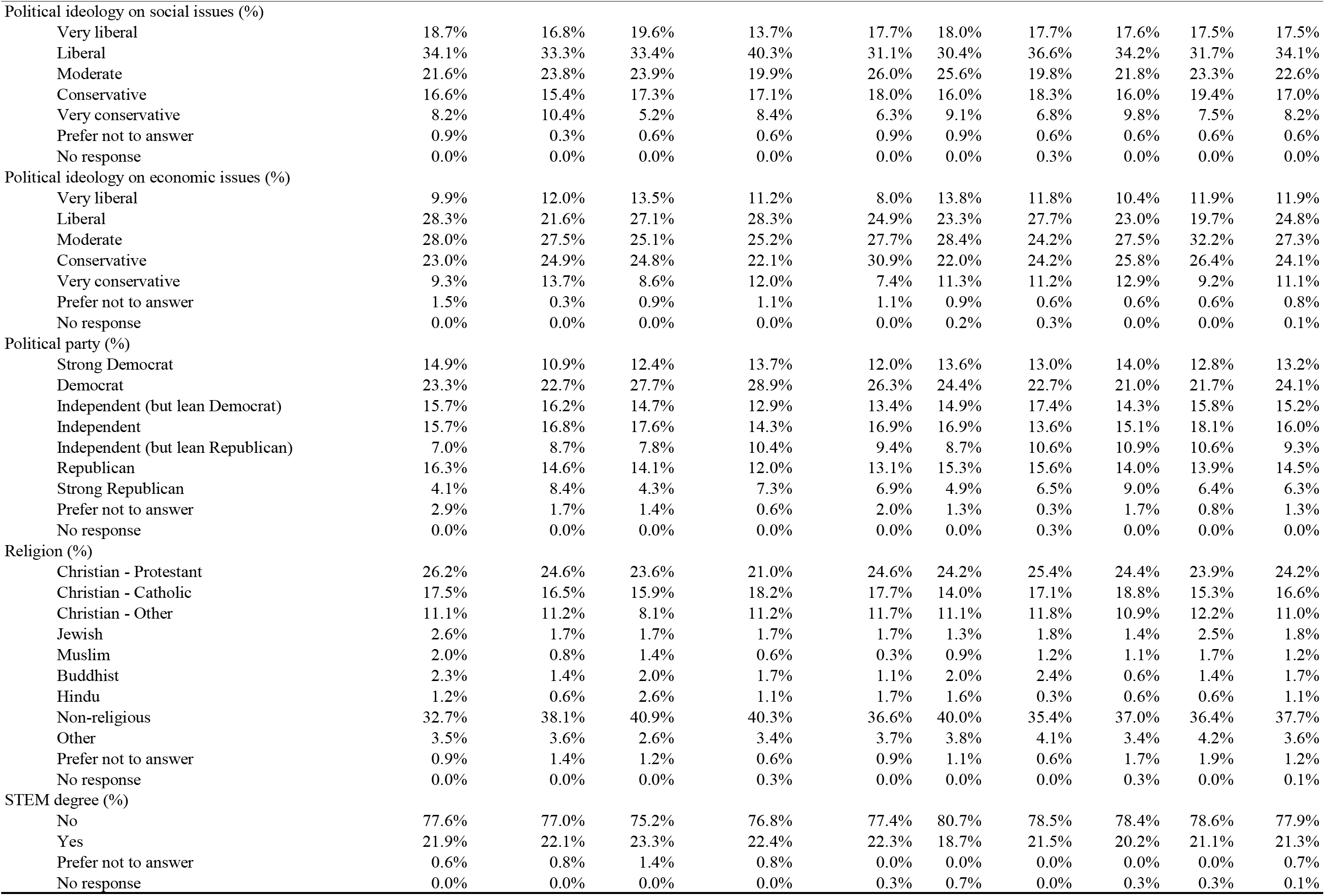
Demographics of lay participants by vignette.

|  | Catheterization<br>Safety<br>Checklist | Best Anti-Hypertensive<br>Drug | Intubation<br>Safety<br>Checklist | Best<br>Corticosteroid<br>Drug | Best<br>Vaccine<br>(first attempt) | Best<br>Vaccine | School<br>Reopening | Ventilator<br>Proning | Masking<br>Rules | All<br>vignettes |
| --- | --- | --- | --- | --- | --- | --- | --- | --- | --- | --- |
| Total N | 343 | 357 | 346 | 357 | 350 | 450 | 339 | 357 | 360 | 2909 |
| Age [Mean (SD)] | 37.9 (12.9) | 38.6 (12.9) | 37.9 (12.4) | 38.0 (12.7) | 36.7 (12.0) | 37.7 (12.6) | 38.7 (13.0) | 39.4 (12.7) | 39.0 (12.8) | 38.4 (12.8) |
| Sex (%) |  |  |  |  |  |  |  |  |  |  |
| Male | 51.3% | 41.5% | 48.1% | 51.5% | 36.6% | 38.4% | 39.2% | 40.9% | 39.7% | 43.6% |
| Female | 47.8% | 58.0% | 51.9% | 48.2% | 63.1% | 60.9% | 60.5% | 58.8% | 60.0% | 55.9% |
| Other | 0.6% | 0.6% | 0.0% | 0.0% | 0.3% | 0.4% | 0.3% | 0.3% | 0.3% | 0.2% |
| Prefer not to answer | 0.3% | 0.0% | 0.0% | 0.3% | 0.0% | 0.2% | 0.0% | 0.0% | 0.0% | 0.2% |
| Race - select all that apply (%) |  |  |  |  |  |  |  |  |  |  |
| Black/African-American | 11.1% | 5.0% | 8.4% | 10.1% | 10.9% | 11.3% | 9.7% | 6.7% | 8.9% | 9.0% |
| Hispanic or Latino | 8.2% | 8.4% | 7.2% | 8.4% | 8.3% | 5.6% | 5.9% | 9.5% | 7.5% | 7.5% |
| White | 72.0% | 78.7% | 71.5% | 72.0% | 70.9% | 72.7% | 77.0% | 77.6% | 75.8% | 74.6% |
| Asian | 12.5% | 8.7% | 15.3% | 12.6% | 12.6% | 13.3% | 8.6% | 7.0% | 7.8% | 10.8% |
| Other | 1.2% | 1.7% | 1.2% | 0.3% | 3.4% | 0.9% | 1.8% | 1.7% | 2.2% | 1.3% |
| Prefer not to answer | 0.9% | 0.6% | 0.0% | 0.6% | 0.3% | 0.9% | 0.6% | 0.3% | 0.3% | 0.5% |
| Education (%) |  |  |  |  |  |  |  |  |  |  |
| Less than high school | 0.6% | 0.8% | 0.3% | 0.3% | 0.6% | 0.2% | 0.3% | 9.8% | 0.8% | 0.4% |
| High school degree | 5.5% | 7.8% | 8.9% | 9.2% | 9.1% | 10.2% | 10.3% | 29.4% | 11.4% | 9.2% |
| Some college | 32.7% | 32.2% | 24.2% | 28.0% | 30.3% | 32.0% | 26.3% | 33.6% | 31.9% | 29.7% |
| Four-year college degree | 37.3% | 35.6% | 39.5% | 35.9% | 37.1% | 35.8% | 37.8% | 3.1% | 30.6% | 35.7% |
| Some graduate school | 4.4% | 3.4% | 4.6% | 4.2% | 4.6% | 5.1% | 4.4% | 23.8% | 4.7% | 4.3% |
| Graduate degree | 19.2% | 19.9% | 22.5% | 22.1% | 18.3% | 16.2% | 20.9% | 0.3% | 20.6% | 20.5% |
| Prefer not to answer | 0.3% | 0.3% | 0.0% | 0.3% | 0.0% | 0.4% | 0.0% | 0.0% | 0.0% | 0.2% |
| Income (%) |  |  |  |  |  |  |  |  |  |  |
| < \$20,000 | 11.1% | 8.4% | 9.2% | 7.6% | 12.0% | 9.3% | 9.4% | 11.2% | 9.7% | 9.5% |
| \$20,000-\$40,000 | 17.8% | 22.1% | 21.6% | 25.8% | 19.7% | 20.2% | 18.9% | 19.0% | 19.7% | 20.7% |
| \$40,000-\$60,000 | 24.5% | 18.8% | 19.0% | 20.2% | 21.4% | 20.4% | 21.2% | 19.9% | 20.8% | 20.6% |
| \$60,000-\$80,000 | 13.7% | 17.4% | 16.1% | 17.9% | 18.6% | 17.8% | 16.5% | 19.3% | 19.2% | 17.3% |
| \$80,000-\$100,000 | 11.4% | 13.7% | 11.0% | 9.5% | 10.6% | 12.2% | 13.3% | 8.4% | 12.2% | 11.5% |
| > \$100,000 | 20.7% | 18.5% | 21.3% | 17.4% | 17.1% | 18.7% | 20.4% | 19.6% | 16.9% | 19.1% |
| Prefer not to answer | 0.9% | 1.1% | 0.9% | 1.4% | 0.3% | 1.3% | 0.3% | 2.5% | 1.4% | 1.2% |
| No response | 0.0% | 0.0% | 0.9% | 0.3% | 0.3% | 0.0% | 0.0% | 0.0% | 0.0% | 0.1% |
| Political Ideology (%) |  |  |  |  |  |  |  |  |  |  |
| Very liberal | 12.2% | 12.6% | 13.0% | 11.2% | 10.6% | 13.1% | 12.7% | 12.0% | 12.8% | 12.5% |
| Liberal | 32.1% | 30.3% | 32.3% | 35.9% | 29.4% | 31.1% | 30.4% | 30.8% | 28.6% | 31.4% |
| Moderate | 29.2% | 25.5% | 28.2% | 26.1% | 31.1% | 27.3% | 27.7% | 24.9% | 28.3% | 27.1% |
| Conservative | 19.8% | 20.2% | 20.7% | 17.1% | 21.7% | 18.7% | 20.9% | 21.3% | 23.6% | 20.2% |
| Very conservative | 5.8% | 10.6% | 5.2% | 9.5% | 6.3% | 8.9% | 7.4% | 9.8% | 5.8% | 7.9% |
| Prefer not to answer | 0.9% | 0.6% | 0.3% | 0.3% | 0.9% | 0.9% | 0.6% | 0.8% | 0.8% | 0.7% |
| No response | 0.0% | 0.3% | 0.3% | 0.0% | 0.0% | 0.0% | 0.3% | 0.3% | 0.0% | 0.1% |
| Political ideology on social issues (%) |  |  |  |  |  |  |  |  |  |  |
| Very liberal | 18.7% | 16.8% | 19.6% | 13.7% | 17.7% | 18.0% | 17.7% | 17.6% | 17.5% | 17.5% |
| Liberal | 34.1% | 33.3% | 33.4% | 40.3% | 31.1% | 30.4% | 36.6% | 34.2% | 31.7% | 34.1% |
| Moderate | 21.6% | 23.8% | 23.9% | 19.9% | 26.0% | 25.6% | 19.8% | 21.8% | 23.3% | 22.6% |
| Conservative | 16.6% | 15.4% | 17.3% | 17.1% | 18.0% | 16.0% | 18.3% | 16.0% | 19.4% | 17.0% |
| Very conservative | 8.2% | 10.4% | 5.2% | 8.4% | 6.3% | 9.1% | 6.8% | 9.8% | 7.5% | 8.2% |
| Prefer not to answer | 0.9% | 0.3% | 0.6% | 0.6% | 0.9% | 0.9% | 0.6% | 0.6% | 0.6% | 0.6% |
| No response | 0.0% | 0.0% | 0.0% | 0.0% | 0.0% | 0.0% | 0.3% | 0.0% | 0.0% | 0.0% |
| Political ideology on economic issues (%) |  |  |  |  |  |  |  |  |  |  |
| Very liberal | 9.9% | 12.0% | 13.5% | 11.2% | 8.0% | 13.8% | 11.8% | 10.4% | 11.9% | 11.9% |
| Liberal | 28.3% | 21.6% | 27.1% | 28.3% | 24.9% | 23.3% | 27.7% | 23.0% | 19.7% | 24.8% |
| Moderate | 28.0% | 27.5% | 25.1% | 25.2% | 27.7% | 28.4% | 24.2% | 27.5% | 32.2% | 27.3% |
| Conservative | 23.0% | 24.9% | 24.8% | 22.1% | 30.9% | 22.0% | 24.2% | 25.8% | 26.4% | 24.1% |
| Very conservative | 9.3% | 13.7% | 8.6% | 12.0% | 7.4% | 11.3% | 11.2% | 12.9% | 9.2% | 11.1% |
| Prefer not to answer | 1.5% | 0.3% | 0.9% | 1.1% | 1.1% | 0.9% | 0.6% | 0.6% | 0.6% | 0.8% |
| No response | 0.0% | 0.0% | 0.0% | 0.0% | 0.0% | 0.2% | 0.3% | 0.0% | 0.0% | 0.1% |
| Political party (%) |  |  |  |  |  |  |  |  |  |  |
| Strong Democrat | 14.9% | 10.9% | 12.4% | 13.7% | 12.0% | 13.6% | 13.0% | 14.0% | 12.8% | 13.2% |
| Democrat | 23.3% | 22.7% | 27.7% | 28.9% | 26.3% | 24.4% | 22.7% | 21.0% | 21.7% | 24.1% |
| Independent (but lean Democrat) | 15.7% | 16.2% | 14.7% | 12.9% | 13.4% | 14.9% | 17.4% | 14.3% | 15.8% | 15.2% |
| Independent | 15.7% | 16.8% | 17.6% | 14.3% | 16.9% | 16.9% | 13.6% | 15.1% | 18.1% | 16.0% |
| Independent (but lean Republican) | 7.0% | 8.7% | 7.8% | 10.4% | 9.4% | 8.7% | 10.6% | 10.9% | 10.6% | 9.3% |
| Republican | 16.3% | 14.6% | 14.1% | 12.0% | 13.1% | 15.3% | 15.6% | 14.0% | 13.9% | 14.5% |
| Strong Republican | 4.1% | 8.4% | 4.3% | 7.3% | 6.9% | 4.9% | 6.5% | 9.0% | 6.4% | 6.3% |
| Prefer not to answer | 2.9% | 1.7% | 1.4% | 0.6% | 2.0% | 1.3% | 0.3% | 1.7% | 0.8% | 1.3% |
| No response | 0.0% | 0.0% | 0.0% | 0.0% | 0.0% | 0.0% | 0.3% | 0.0% | 0.0% | 0.0% |
| Religion (%) |  |  |  |  |  |  |  |  |  |  |
| Christian - Protestant | 26.2% | 24.6% | 23.6% | 21.0% | 24.6% | 24.2% | 25.4% | 24.4% | 23.9% | 24.2% |
| Christian - Catholic | 17.5% | 16.5% | 15.9% | 18.2% | 17.7% | 14.0% | 17.1% | 18.8% | 15.3% | 16.6% |
| Christian - Other | 11.1% | 11.2% | 8.1% | 11.2% | 11.7% | 11.1% | 11.8% | 10.9% | 12.2% | 11.0% |
| Jewish | 2.6% | 1.7% | 1.7% | 1.7% | 1.7% | 1.3% | 1.8% | 1.4% | 2.5% | 1.8% |
| Muslim | 2.0% | 0.8% | 1.4% | 0.6% | 0.3% | 0.9% | 1.2% | 1.1% | 1.7% | 1.2% |
| Buddhist | 2.3% | 1.4% | 2.0% | 1.7% | 1.1% | 2.0% | 2.4% | 0.6% | 1.4% | 1.7% |
| Hindu | 1.2% | 0.6% | 2.6% | 1.1% | 1.7% | 1.6% | 0.3% | 0.6% | 0.6% | 1.1% |
| Non-religious | 32.7% | 38.1% | 40.9% | 40.3% | 36.6% | 40.0% | 35.4% | 37.0% | 36.4% | 37.7% |
| Other | 3.5% | 3.6% | 2.6% | 3.4% | 3.7% | 3.8% | 4.1% | 3.4% | 4.2% | 3.6% |
| Prefer not to answer | 0.9% | 1.4% | 1.2% | 0.6% | 0.9% | 1.1% | 0.6% | 1.7% | 1.9% | 1.2% |
| No response | 0.0% | 0.0% | 0.0% | 0.3% | 0.0% | 0.0% | 0.0% | 0.3% | 0.0% | 0.1% |
| STEM degree (%) |  |  |  |  |  |  |  |  |  |  |
| No | 77.6% | 77.0% | 75.2% | 76.8% | 77.4% | 80.7% | 78.5% | 78.4% | 78.6% | 77.9% |
| Yes | 21.9% | 22.1% | 23.3% | 22.4% | 22.3% | 18.7% | 21.5% | 20.2% | 21.1% | 21.3% |
| Prefer not to answer | 0.6% | 0.8% | 1.4% | 0.8% | 0.0% | 0.0% | 0.0% | 0.0% | 0.0% | 0.7% |
| No response | 0.0% | 0.0% | 0.0% | 0.0% | 0.3% | 0.7% | 0.0% | 0.3% | 0.3% | 0.1% |

#### Clinicians

There were 2,149 clinician responses across all vignettes. In the clinician samples, survey responses were anonymous, so we could not restrict participation based on our previous studies so some participants who completed the Intubation Safety Checklist, Best Corticosteroid Drug, and Masking Rules vignettes may have also completed the Best Vaccine vignette. For this reason, demographics are reported separately by vignette in Table S5. Across vignettes, a majority of clinicians were female. Over 50% of participants in the sample were registered nurses, followed by physicians and physician assistants. Over 50% of participants in the sample reported that they had been in the medical field for over 10 years. The clinicians reported that they had received training in research methods and statistics via an average of 1.5 of the sources we listed, and that they engaged in an average of 2.5 research methods and statistics activities. Most clinicians reported being somewhat to moderately comfortable with research methods and statistics.

**Table S5.** Demographics of clinicians by vignette.

|  | Intubation<br>Safety<br>Checklist | Best<br>Corticosteroid<br>Drug | Masking<br>Rules | Best<br>Vaccine |
| --- | --- | --- | --- | --- |
| Total N | 271 | 275 | 349 | 1254 |
| Sex (%) |  |  |  |  |
| Male | 18.1% | 22.5% | 18.1% | 18.7% |
| Female | 81.9% | 77.1% | 81.4% | 81.2% |
| Other | 0.0% | 0.4% | 0.6% | 0.2% |
| Source of research methods/statistics training - select all that apply (%) |  |  |  |  |
| Undergraduate coursework | 48.7% | 49.5% | 48.7% | 47.4% |
| Professional school instruction | 40.2% | 31.3% | 34.4% | 34.4% |
| Postgraduate coursework | 26.2% | 20.7% | 22.1% | 21.1% |
| CME/CEU courses | 27.7% | 25.1% | 24.1% | 25.8% |
| Self-instruction via peer-reviewed literature | 19.2% | 15.6% | 17.2% | 21.3% |
| Other | 7.0% | 4.0% | 3.2% | 3.9% |
| Total number of research methods/statistics training [mean (SD)] | 1.69 (1.22) | 1.46 (1.02) | 1.50 (1.13) | 1.54 (1.16) |
| Comfort with research methods/statistics (%) |  |  |  |  |
| Not at all | 8.9% | 12.7% | 10.9% | 11.1% |
| Somewhat | 37.6% | 44.4% | 45.8% | 46.6% |
| Moderately | 39.5% | 32.0% | 32.7% | 30.8% |
| Very | 11.8% | 9.1% | 8.9% | 9.9% |
| Extremely | 2.2% | 1.8% | 1.7% | 1.7% |
| Research methods/statistics activities - select all that apply (%) |  |  |  |  |
| Read results of RCT in peer-reviewed journal article | 81.2% | 75.3% | 71.9% | 71.2% |
| Changed typical prescription/recommendation after personally reading results of RCT in peer-reviewed journal article | 41.0% | 33.1% | 33.0% | 39.8% |
| Published scientific paper in peer-reviewed journal | 13.3% | 12.4% | 9.7% | 12.0% |
| Conducted or worked on a team conducting an RCT | 18.5% | 20.0% | 19.2% | 17.1% |
| Took a course/class in statistics, biostatistics, research methods | 73.1% | 69.8% | 69.1% | 68.5% |
| Analyzed data for statistical significance outside of course requirement | 23.6% | 21.8% | 19.2% | 21.1% |
| Used statistical software | 12.2% | 11.6% | 11.5% | 9.3% |
| Total number of research methods/statistics activities [mean (SD)] | 2.63 (1.69) | 2.44 (1.71) | 2.34 (1.66) | 2.39 (1.72) |
| Currently involved in research (%) | 10.7% | 9.1% | 9.7% | 9.6% |
| Position (%) |  |  |  |  |
| Doctor | 14.8% | 14.5% | 12.6% | 15.7% |
| Physician Assistant | 12.5% | 6.9% | 9.5% | 7.7% |
| Nurse Practitioner | 6.3% | 2.5% | 4.3% | 4.7% |
| Nurse (RN) | 51.3% | 57.1% | 55.6% | 52.8% |
| Nurse (LPN) | 6.3% | 9.5% | 8.0% | 15.6% |
| Nurse (Other) | 1.8% | 1.1% | 1.4% | 0.6% |
| Genetic Counselor | 0.0% | 0.0% | 0.0% | 0.0% |
| Non-prescribing clinician or staff without clinical credential | 0.0% | 0.0% | 0.0% | 0.0% |
| Medical student | 5.2% | 5.5% | 4.6% | 0.1% |
| Faculty or Professor | 0.4% | 0.7% | 0.3% | 0.3% |
| Other | 1.5% | 2.2% | 3.7% | 2.6% |
| Years in medical field (%) |  |  |  |  |
| < 1 year | 2.6% | 2.9% | 3.2% | 2.8% |
| 1-2 years | 6.3% | 5.5% | 6.0% | 5.8% |
| 3-5 years | 15.1% | 11.3% | 12.6% | 13.6% |
| 6-10 years | 16.6% | 14.2% | 15.8% | 15.8% |
| > 10 years | 59.4% | 66.2% | 62.5% | 62.0% |

*Note.* Reported here are the demographics of the clinicians who saw the Intubation Safety Checklist, Best Corticosteroid Drug, or Masking Rules vignette first (responses to the Best Vaccine vignette were collected at a different time). All clinicians who participated in this study completed all vignettes but in randomized order. In the main text, we only analyze responses to the first vignette so we report demographics similarly here.

### Results presented in main text

In Table S6A-C, we present the descriptive and inferential results for all vignettes discussed in the main text.

**Table S6A.** Descriptive and inferential results of ratings and rankings of interventions and experiment for all vignettes.

| Descriptive Results |  |  |  |  | Inferential Results |  |
| --- | --- | --- | --- | --- | --- | --- |
| Vignette | Variable | Mean (SD) | % Ranking Best | % Ranking Worst | Test Description | Test Outcome |
| Lay Sentiments About Healthcare Experimentation |  |  |  |  |  |  |
| Catheterization Safety Checklist<br>(*n* = 343 laypeople) | | | | | A/B Effect | $t(342) = 9.74^{***}, d = 0.69 \pm .16$ |
| | | | | | Mean(A,B) > AB | 58% ± 5% |
| | A | 3.77 (1.12) | 27% | 32% | Reverse A/B effect | $t(342) = -9.74^{***}, d = -0.69 \pm .16$ |
| | B | 4.03 (1.09) | 42% | 21% | AB > Mean(A,B) | 27% ± 4% |
| | AB | 3.09 (1.40) | 32% | 48% | Experiment Aversion | $t(342) = 3.70^{***}, d = 0.25 \pm .14$ |
| | Mean(A,B) | 3.90 (0.84) | - | - | Min(A,B) > AB | 41% ± 5% |
| | Min(A,B) | 3.42 (1.16) | - | - | Experiment Appreciation | $t(342) = -14.61^{***}, d = -1.13 \pm .20$ |
| | Max(A,B) | 4.39 (0.81) | - | - | AB > Max(A,B) | 15% ± 3% |
| | | | | | Experiment Rejection<br>(A,B = 3,4,5; AB = 1,2) | 28% ± 5% |
| | | | | | Experiment Endorsement<br>(AB = 4,5; A,B = 1,2,3) | 3% ± 1% |
| Best Anti-Hypertensive Drug<br>(*n* = 357 laypeople) | | | | | A/B Effect | $t(356) = 6.68^{***}, d = 0.52 \pm .16$ |
| | | | | | Mean(A,B) > AB | 47% ± 5% |
| | A | 3.87 (1.00) | 25% | 27% | Reverse A/B effect | $t(356) = -6.68^{***}, d = -0.52 \pm .16$ |
| | B | 3.89 (0.99) | 25% | 28% | AB > Mean(A,B) | 31% ± 5% |
| | AB | 3.24 (1.47) | 50% | 45% | Experiment Aversion | $t(356) = 5.96^{***}, d = 0.46 \pm .16$ |
| | Mean(A,B) | 3.88 (0.95) | - | - | Min(A,B) > AB | 44% ± 5% |
| | Min(A,B) | 3.82 (1.03) | - | - | Experiment Appreciation | $t(356) = -7.26^{***}, d = -0.57 \pm .17$ |
| | Max(A,B) | 3.94 (0.95) | - | - | AB > Max(A,B) | 29% ± 4% |
| | | | | | Experiment Rejection<br>(A,B = 3,4,5; AB = 1,2) | 34% ± 5% |
| | | | | | Experiment Endorsement<br>(AB = 4,5; A,B = 1,2,3) | 18% ± 4% |

*Note.* The A/B Effect refers to the difference between the average rating of the two interventions and the rating of the A/B test. Mean(A,B) > AB is the percentage of people whose average intervention rating was higher than their rating of the A/B test. The Reverse A/B Effect refers to difference between the rating of the A/B test and the average rating of the two interventions. AB > Mean(A,B) is the percentage of people who rating of the A/B test was higher than their average intervention rating. Experiment Aversion refers to the difference between the rating of the A/B test and the lowest-rated intervention. Min(A,B) > AB is the percentage of people whose lowest-rated intervention is rated higher than their rating of the A/B test. Experiment Appreciation refers to the difference between the rating of the highest-rated intervention and the rating of the A/B test. AB > Max(A,B) is the percentage of people whose rating of the A/B test is higher than the rating of their highest-rated intervention. Experiment Rejection is the percentage of people who rated interventions A and B as "neither inappropriate nor appropriate" or more appropriate while rating the A/B test as "very" or "somewhat" inappropriate. Experiment Endorsement is the percentage of people who rated the A/B test as "very" or "somewhat" appropriate while rating interventions A and B as "neither inappropriate nor appropriate" or less appropriate.
\* $p < .05$
\*\* $p < .01$
\*\*\* $p < .001$

**Table S6B.**
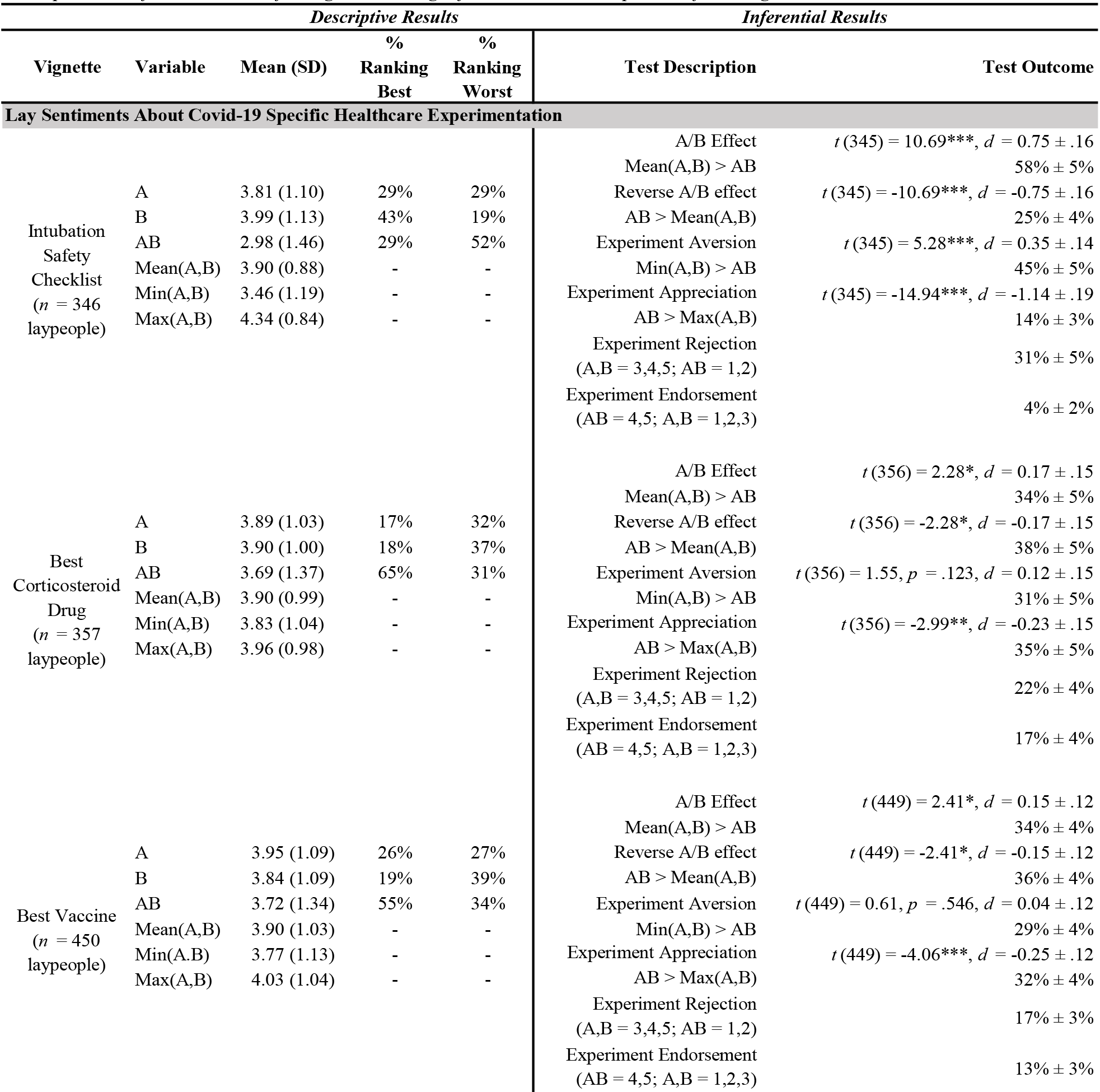
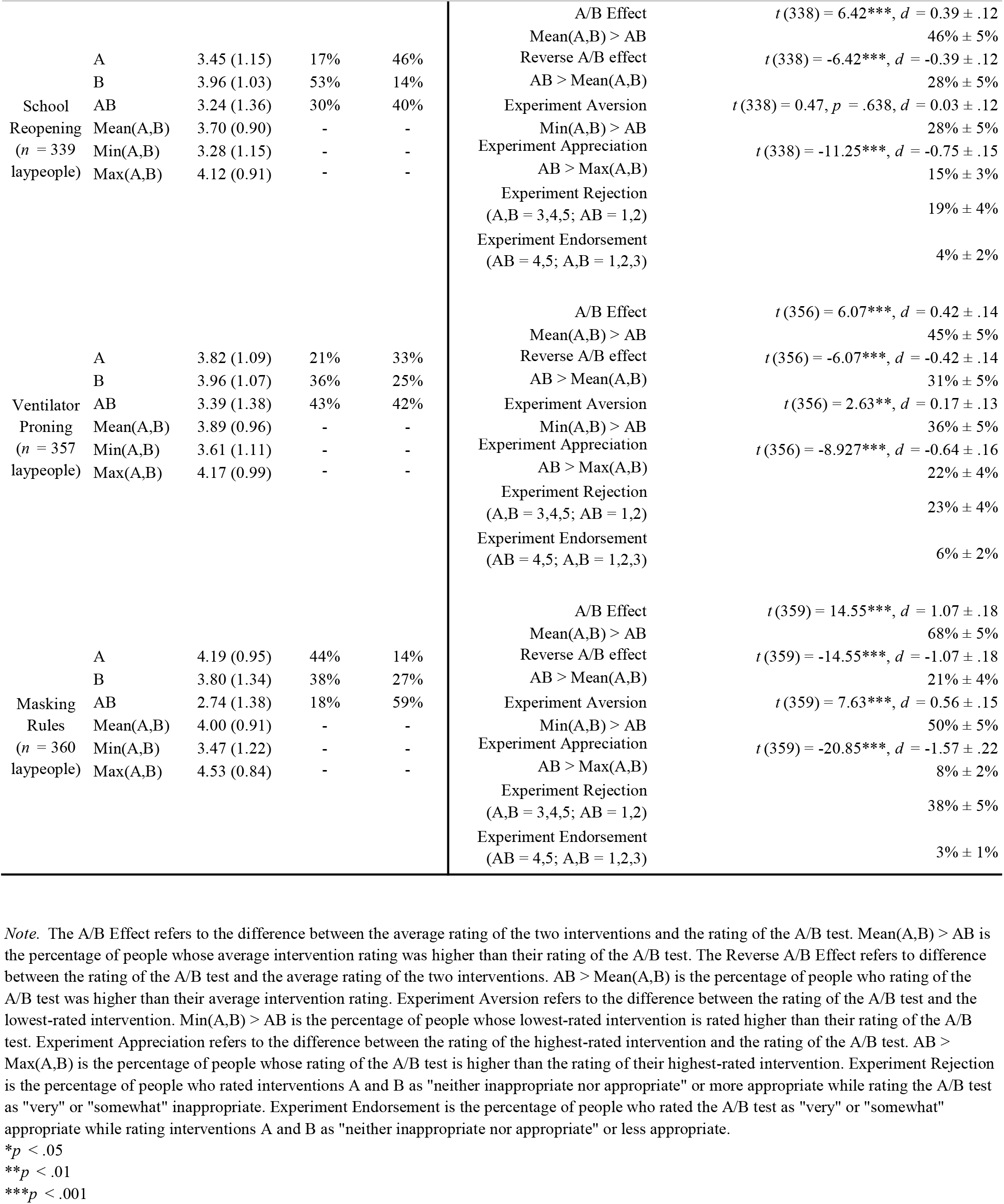
Descriptive and inferential results of ratings and rankings of interventions and experiment for all vignettes.

**Table S6C.**
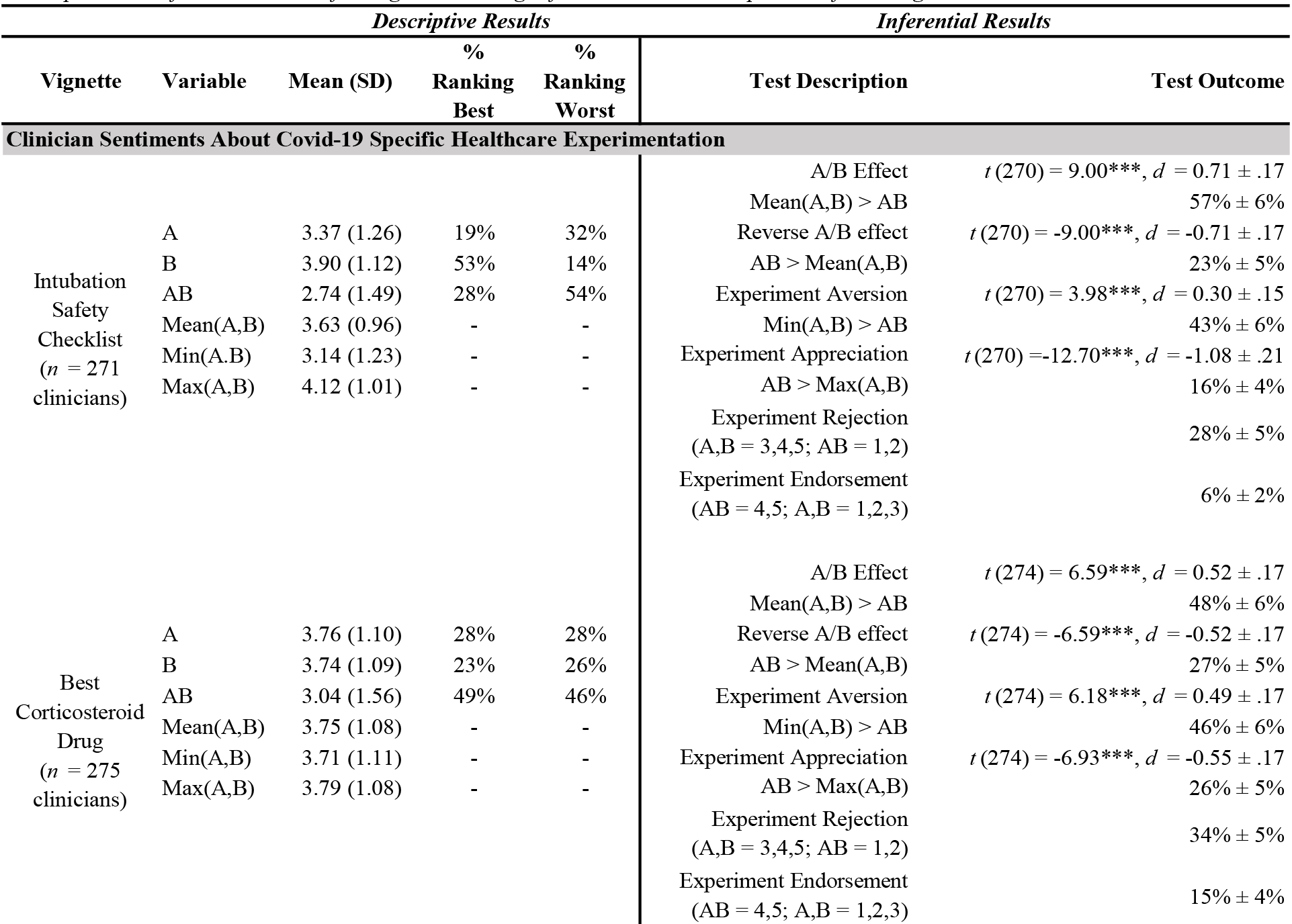
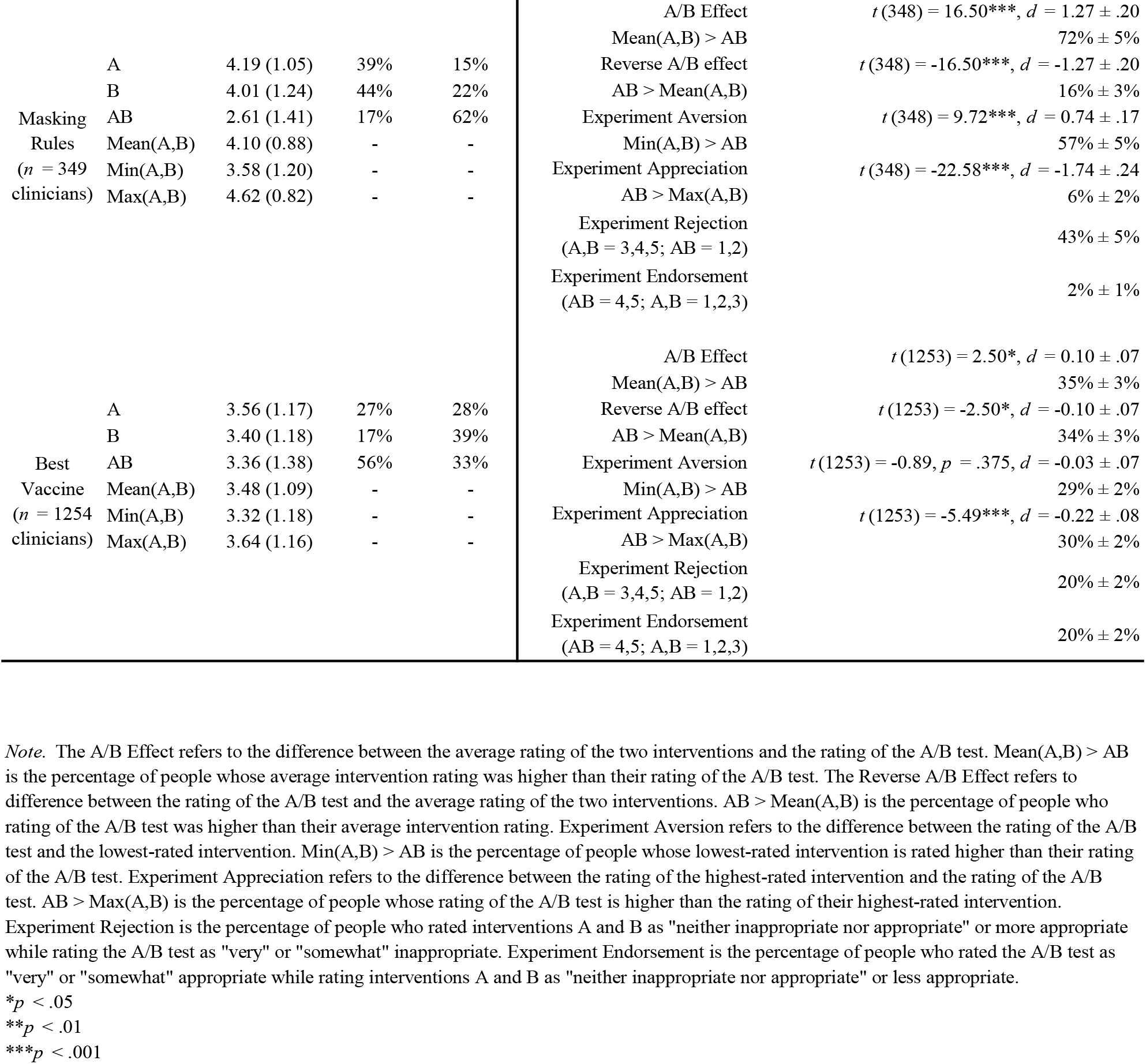
Descriptive and inferential results of ratings and rankings of interventions and experiment for all vignettes.

#### Comparisons to previously published work

To compare these results to our previous findings reporting sentiments about experiments, as we do in the main text, please refer to Heck et al. (2020)^2^. For example, in the Results section “Lay Sentiments About Healthcare Experimentation,” we say, “these levels of experiment aversion near the height of the pandemic were slightly (but not significantly) higher than those we observed among similar laypeople in 2019 (41% ± 5% in 2020 vs. 37% ± 6% in 2019 for Catheterization Safety Checklist, *p* = .31 ; 44% ± 5% in 2020 vs. 40% ± 6% in 2019 for Best Anti-Hypertensive Drug, *p* = .32).” We extracted the percentage of participants who were experiment averse in 2019 from Heck et al. (2020)^2^. We then performed a two-sample z-test for proportions to compare the 2019 and 2020 proportions. As noted in the main text, we did not find a significant difference between the percentage of people who were experiment averse in 2019 and the percentage of people who were experiment averse in the current studies which took place in 2020 and 2021 (Catheterization Safety Checklist: χ^2^(1) = 1.034, *p* = .309, Anti-Hypertensive Drug: χ^2^(1) = 0.998, *p* = .318).

### Results not presented in the main text

#### Results of Best Vaccine vignette (initial ambiguous version)

The only vignette which showed no A/B Effect was the initial ambiguous version of Best Vaccine (see Table S6D). The two versions of Best Vaccine both presented a public health official’s decision to either distribute an mRNA-based vaccine to every county in their state, distribute an inactivated-virus vaccine to every county, or run an experiment in which counties are randomized to receive one of the two vaccine types. However, in version 1, the wording unintentionally implied that residents could choose their vaccine (by going elsewhere) if they did not wish to be subject to the official’s decision (including intervention implementation or A/B test), while in version 2 we eliminated this possible interpretation; we suspect this had the effect of making the experiment condition in version 1 less aversive, since people could effectively opt-out of it, and our goal in this research is to study pragmatic, real-world situations in which avoiding randomization is typically not a realistic option.

**Table S6D.**
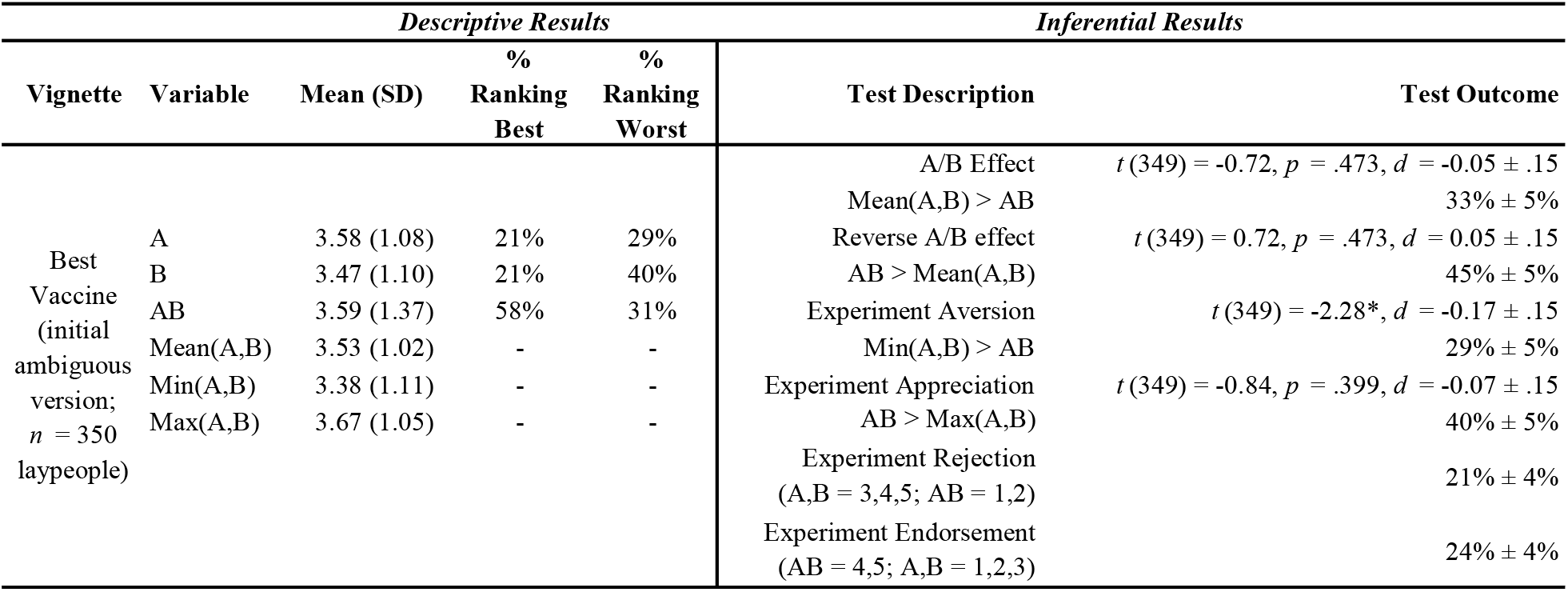
Descriptive and inferential results of ratings and rankings of interventions and experiment for all vignettes.

| *Descriptive Results* |  |  |  |  | *Inferential Results* |  |
| --- | --- | --- | --- | --- | --- | --- |
| Vignette | Variable | Mean (SD) | %<br>Ranking<br>Best | %<br>Ranking<br>Worst | Test Description | Test Outcome |
| Best<br>Vaccine<br>(initial<br>ambiguous<br>version;<br>$n = 350$<br>laypeople) | A | 3.58 (1.08) | 21% | 29% | A/B Effect<br>Mean(A,B) > AB | $t(349) = -0.72, p = .473, d = -0.05 \pm .15$<br>33% ± 5% |
| | B | 3.47 (1.10) | 21% | 40% | Reverse A/B effect<br>AB > Mean(A,B) | $t(349) = 0.72, p = .473, d = 0.05 \pm .15$<br>45% ± 5% |
| | AB | 3.59 (1.37) | 58% | 31% | Experiment Aversion<br>Min(A,B) > AB | $t(349) = -2.28^*, d = -0.17 \pm .15$<br>29% ± 5% |
| | Mean(A,B) | 3.53 (1.02) | - | - | Experiment Appreciation<br>AB > Max(A,B) | $t(349) = -0.84, p = .399, d = -0.07 \pm .15$<br>40% ± 5% |
| | Min(A,B) | 3.38 (1.11) | - | - | Experiment Rejection<br>(A,B = 3,4,5; AB = 1,2) | 21% ± 4% |
| | Max(A,B) | 3.67 (1.05) | - | - | Experiment Endorsement<br>(AB = 4,5; A,B = 1,2,3) | 24% ± 4% |

#### Order effect in clinician study

For the clinician study of the Catheterization Safety Checklist, Best Anti-Hypertensive Drug, and Masking Rules vignettes, participants were randomly assigned to one of these three vignettes and then completed the remaining two vignettes in random order. For consistency with the rest of this project and with our previous approach (Meyer et al., 2019)^1^, we analyze data from this study as a between-subjects design where we only consider the first vignette that every participant completed.

While conducting an interim analysis on the data for this study, we observed an intriguing and unexpected order effect of presentation.

For the first 601 complete responses we received, we observed an effect of presentation order on participants’ appropriateness ratings of the A/B test condition within the Best Anti-Hypertensive Drug vignette. Participants who received the Best Anti-Hypertensive Drug vignette first rated the A/B test an average of 2.95 (SD = 1.57), participants who received this vignette second rated the A/B test an average of 3.48 (SD = 1.39), and participants who received this vignette last rated the A/B test an average of 3.78 (SD = 1.41). This suggests that participants who read about other policies and A/B tests before considering the Best Anti-Hypertensive Drug vignette found the A/B test in the Best Anti-Hypertensive Drug vignette to be less objectionable than participants who received this vignette earlier in the survey. The relationship between presentation order (1, 2, or 3) and appropriateness rating of the A/B test was *r* = .23. This order effect did not emerge for the other two vignettes or for ratings of either intervention (A or B).

After observing this order effect but before examining any additional data, we preregistered this order effect with the goal of replicating it in an independent sample. 294 new participants completed the study after this interim analysis, and we analyzed the data from this sample independently from the sample that generated the order effect. Table S7 displays ratings of the A/B condition within each scenario grouped by the order in which participants received them. The order effect observed with the Best Anti-Hypertensive Drug A/B test condition replicated (*r* = .15), as did the absence of any similar order effect for the other conditions.

**Table S7.** Ratings of A/B test in Clinician Sample.

| **Exploratory Sample (N = 601)** | **Best Corticosteroid Drug** | **Intubation Safety Checklist** | **Masking Rules** |
| --- | --- | --- | --- |
|  | **A/B Rating (SD)** | **A/B Rating (SD)** | **A/B Rating (SD)** |
| Target Scenario First | 2.95 (1.57) | 2.79 (1.49) | 2.63 (1.43) |
| Target Scenario Second | 3.48 (1.39) | 2.53 (1.35) | 2.66 (1.44) |
| Target Scenario Last | 3.78 (1.41) | 2.78 (1.38) | 2.57 (1.29) |

| **Confirmatory Sample (N = 294)** | **Best Corticosteroid Drug** | **Intubation Safety Checklist** | **Masking Rules** |
| --- | --- | --- | --- |
|  | **A/B Rating (SD)** | **A/B Rating (SD)** | **A/B Rating (SD)** |
| Target Scenario First | 3.22 (1.54) | 2.63 (1.50) | 2.58 (1.38) |
| Target Scenario Second | 3.49 (1.51) | 2.76 (1.39) | 2.38 (1.42) |
| Target Scenario Last | 3.77 (1.33) | 2.69 (1.15) | 2.51 (1.38) |

#### Heterogeneity in experiment aversion

In both the lay participant sample and the clinician sample, associations between demographic variables, including educational attainment, having a degree in a STEM field, years of experience in the medical field, and role in the healthcare system, and sentiment about experimentation (e.g., A/B effect, experiment aversion, experiment appreciation) are consistently small (*r* < |.13|, therefore explaining less than 2% of the variance; Tables S8–11).

**Table S8.** Correlations between lay participant characteristics and sentiments about experiments.

|  | Size of A/B effect |  | A/B effect |  | Size of experiment aversion |  | Experiment aversion |  | Experiment rejection |  | Size of experiment appreciation |  | Experiment appreciation |  | Experiment endorsement |  |
| --- | --- | --- | --- | --- | --- | --- | --- | --- | --- | --- | --- | --- | --- | --- | --- | --- |
|  | *r* | *p* | *r* | *p* | *r* | *p* | *r* | *p* | *r* | *p* | *r* | *p* | *r* | *p* | *r* | *p* |
| Age | -0.008 | 0.662 | -0.020 | 0.286 | -0.020 | 0.270 | -0.038 | 0.043 | -0.046 | 0.012 | -0.004 | 0.809 | -0.016 | 0.389 | -0.033 | 0.073 |
| Sex<br>(1 = male, 2 = female) | 0.068 | <.001 | 0.048 | 0.010 | 0.067 | <.001 | 0.039 | 0.035 | 0.059 | 0.002 | -0.064 | <.001 | -0.071 | <.001 | -0.036 | 0.053 |
| Race<br>(0 = all other, 1 = Nonhispanic White) | -0.004 | 0.814 | -0.017 | 0.360 | -0.001 | 0.945 | -0.016 | 0.388 | 0.003 | 0.867 | 0.007 | 0.706 | 0.001 | 0.937 | -0.012 | 0.533 |
| Education | 0.047 | 0.011 | 0.033 | 0.075 | 0.049 | 0.008 | 0.051 | 0.006 | 0.029 | 0.114 | -0.042 | 0.024 | -0.023 | 0.216 | -0.019 | 0.298 |
| Income | 0.020 | 0.293 | 0.005 | 0.787 | 0.020 | 0.273 | 0.011 | 0.571 | 0.005 | 0.777 | -0.017 | 0.353 | -0.025 | 0.184 | -0.026 | 0.158 |
| Political Ideology<br>(1 = Very Liberal, 5 = Very Conservative) | -0.114 | <.0001 | -0.087 | <.0001 | -0.118 | <.0001 | -0.101 | <.0001 | -0.091 | <.0001 | 0.101 | <.0001 | 0.043 | 0.022 | 0.045 | 0.015 |
| Political Ideology (Social)<br>(1 = Very Liberal, 5 = Very Conservative) | -0.123 | <.0001 | -0.099 | <.0001 | -0.128 | <.0001 | -0.118 | <.0001 | -0.106 | <.0001 | 0.109 | <.0001 | 0.039 | 0.036 | 0.052 | 0.005 |
| Political Ideology (Economic)<br>(1 = Very Liberal, 5 = Very Conservative) | -0.094 | <.0001 | -0.065 | <.001 | -0.095 | <.0001 | -0.082 | <.0001 | -0.073 | <.0001 | 0.085 | <.0001 | 0.046 | 0.013 | 0.040 | 0.031 |
| Political Party<br>(1 = Strong Democrat, 7 = Strong Republican) | -0.096 | <.0001 | -0.073 | <.0001 | -0.098 | <.0001 | -0.075 | <.0001 | -0.075 | <.0001 | 0.087 | <.0001 | 0.037 | 0.050 | 0.035 | 0.063 |
| Conservatism<br>(mean of z-scored Political Ideology, Political Ideology (Social), Political Ideology (Economic), and Political Party) | -0.117 | <.0001 | -0.089 | <.0001 | -0.121 | <.0001 | -0.103 | <.0001 | -0.095 | <.0001 | 0.105 | <.0001 | 0.045 | 0.015 | 0.047 | 0.012 |
| Non-religious<br>(0 = Religious (any religion), 1 = Non-religious) | 0.051 | 0.007 | 0.027 | 0.150 | 0.061 | <.001 | 0.049 | 0.009 | 0.046 | 0.015 | -0.036 | 0.053 | -0.013 | 0.496 | -0.021 | 0.266 |
| STEM degree<br>(0 = no, 1 = yes) | 0.023 | 0.208 | 0.016 | 0.399 | 0.027 | 0.154 | 0.026 | 0.157 | 0.027 | 0.142 | -0.019 | 0.318 | 0.016 | 0.403 | 0.024 | 0.205 |

**Table S9.**
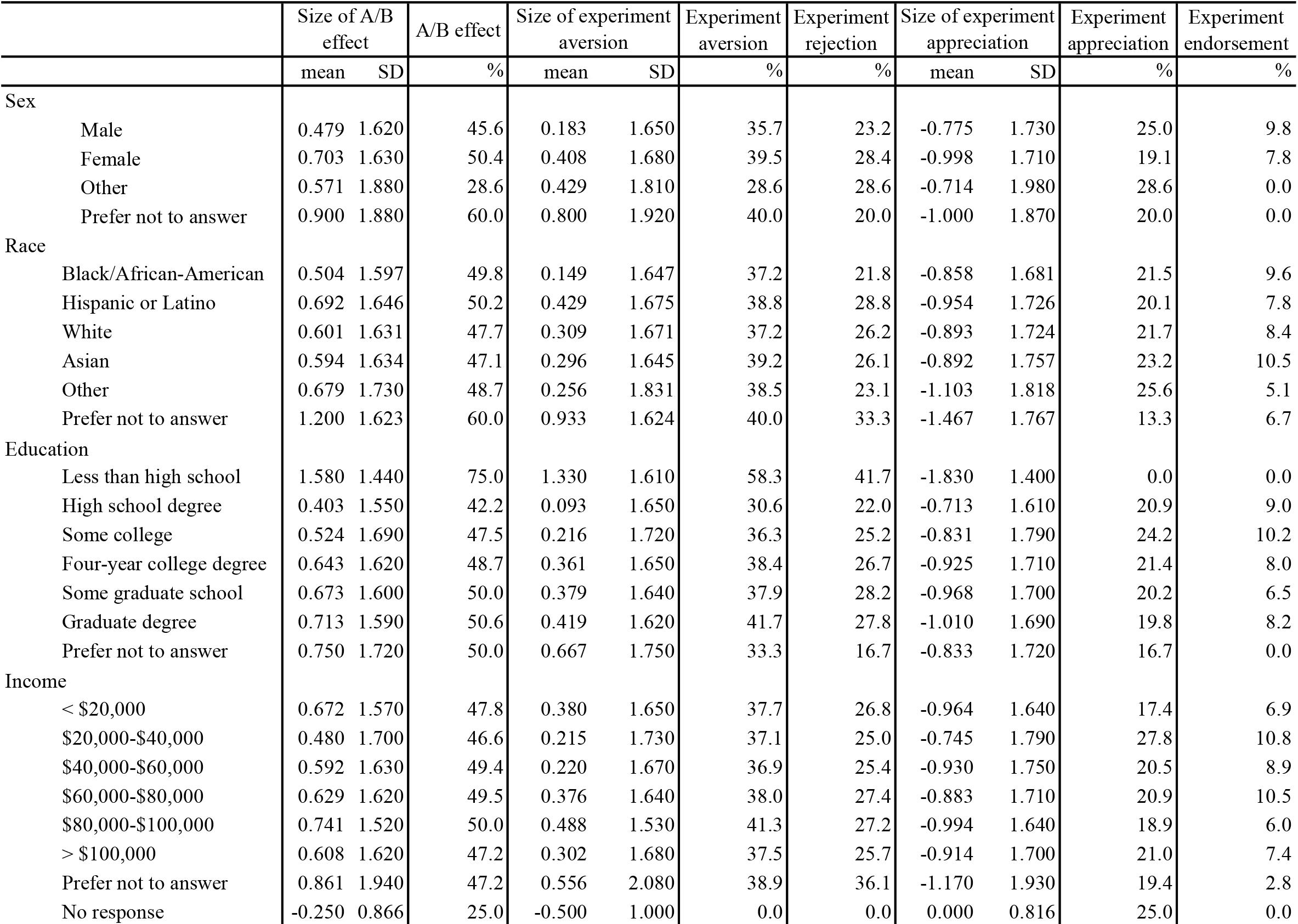
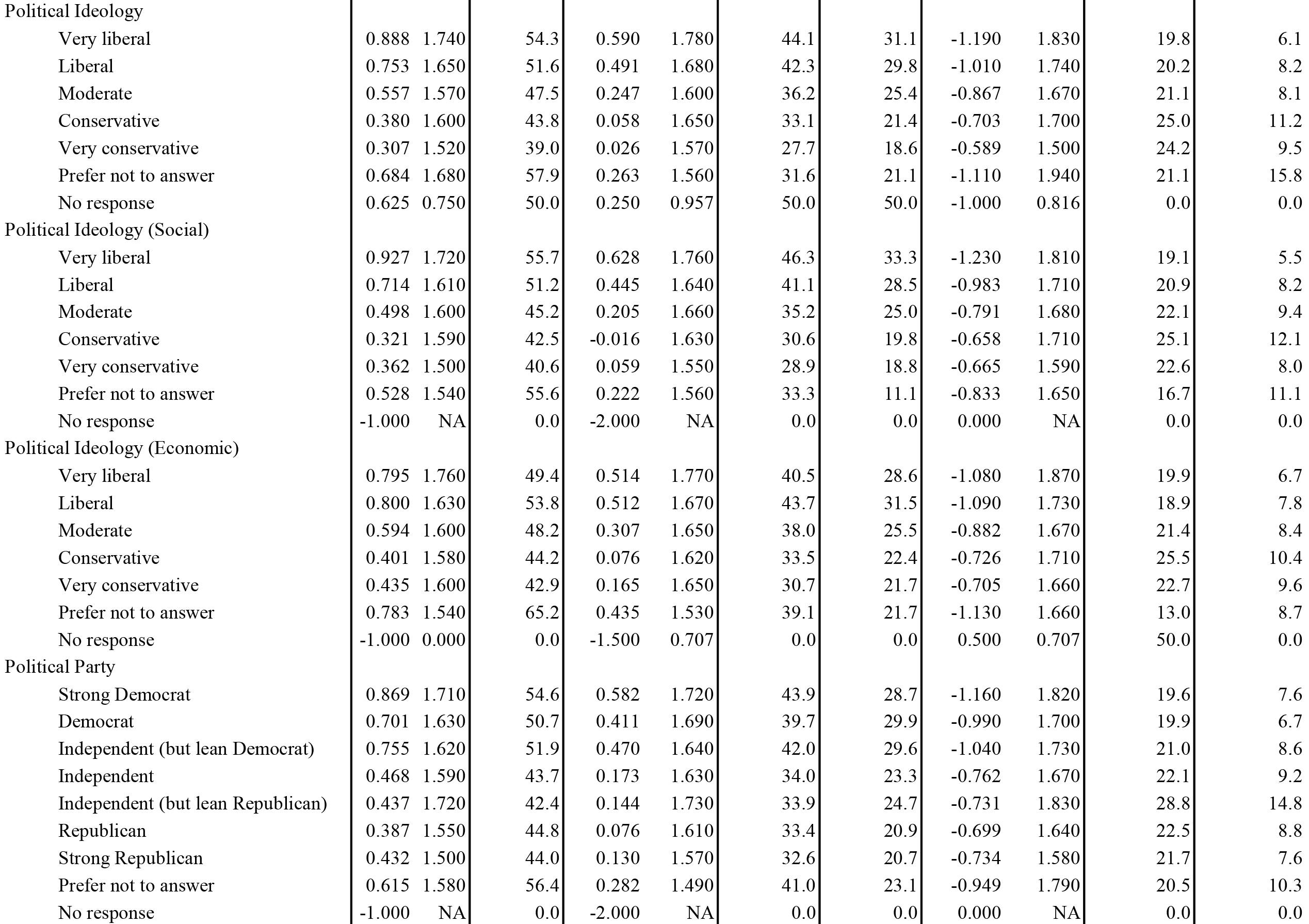
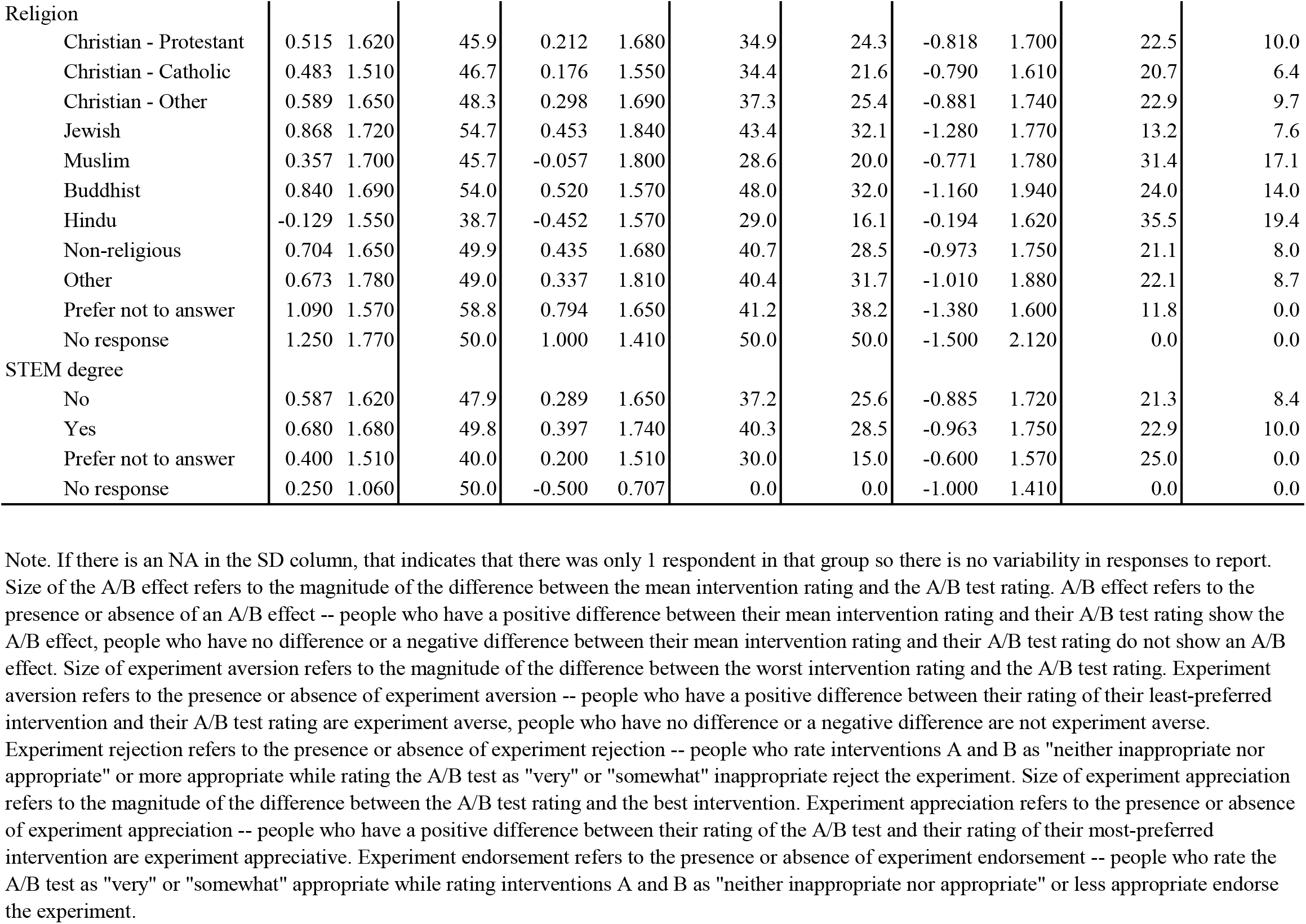
Means and percentages of sentiments about experiments by demographic variable in lay participants.

|  | Size of A/B effect |  | A/B effect | Size of experiment aversion |  | Experiment aversion | Experiment rejection | Size of experiment appreciation |  | Experiment appreciation | Experiment endorsement |
| --- | --- | --- | --- | --- | --- | --- | --- | --- | --- | --- | --- |
|  | mean | SD | % | mean | SD | % | % | mean | SD | % | % |
| Sex |  |  |  |  |  |  |  |  |  |  |  |
| Male | 0.479 | 1.620 | 45.6 | 0.183 | 1.650 | 35.7 | 23.2 | -0.775 | 1.730 | 25.0 | 9.8 |
| Female | 0.703 | 1.630 | 50.4 | 0.408 | 1.680 | 39.5 | 28.4 | -0.998 | 1.710 | 19.1 | 7.8 |
| Other | 0.571 | 1.880 | 28.6 | 0.429 | 1.810 | 28.6 | 28.6 | -0.714 | 1.980 | 28.6 | 0.0 |
| Prefer not to answer | 0.900 | 1.880 | 60.0 | 0.800 | 1.920 | 40.0 | 20.0 | -1.000 | 1.870 | 20.0 | 0.0 |
| Race |  |  |  |  |  |  |  |  |  |  |  |
| Black/African-American | 0.504 | 1.597 | 49.8 | 0.149 | 1.647 | 37.2 | 21.8 | -0.858 | 1.681 | 21.5 | 9.6 |
| Hispanic or Latino | 0.692 | 1.646 | 50.2 | 0.429 | 1.675 | 38.8 | 28.8 | -0.954 | 1.726 | 20.1 | 7.8 |
| White | 0.601 | 1.631 | 47.7 | 0.309 | 1.671 | 37.2 | 26.2 | -0.893 | 1.724 | 21.7 | 8.4 |
| Asian | 0.594 | 1.634 | 47.1 | 0.296 | 1.645 | 39.2 | 26.1 | -0.892 | 1.757 | 23.2 | 10.5 |
| Other | 0.679 | 1.730 | 48.7 | 0.256 | 1.831 | 38.5 | 23.1 | -1.103 | 1.818 | 25.6 | 5.1 |
| Prefer not to answer | 1.200 | 1.623 | 60.0 | 0.933 | 1.624 | 40.0 | 33.3 | -1.467 | 1.767 | 13.3 | 6.7 |
| Education |  |  |  |  |  |  |  |  |  |  |  |
| Less than high school | 1.580 | 1.440 | 75.0 | 1.330 | 1.610 | 58.3 | 41.7 | -1.830 | 1.400 | 0.0 | 0.0 |
| High school degree | 0.403 | 1.550 | 42.2 | 0.093 | 1.650 | 30.6 | 22.0 | -0.713 | 1.610 | 20.9 | 9.0 |
| Some college | 0.524 | 1.690 | 47.5 | 0.216 | 1.720 | 36.3 | 25.2 | -0.831 | 1.790 | 24.2 | 10.2 |
| Four-year college degree | 0.643 | 1.620 | 48.7 | 0.361 | 1.650 | 38.4 | 26.7 | -0.925 | 1.710 | 21.4 | 8.0 |
| Some graduate school | 0.673 | 1.600 | 50.0 | 0.379 | 1.640 | 37.9 | 28.2 | -0.968 | 1.700 | 20.2 | 6.5 |
| Graduate degree | 0.713 | 1.590 | 50.6 | 0.419 | 1.620 | 41.7 | 27.8 | -1.010 | 1.690 | 19.8 | 8.2 |
| Prefer not to answer | 0.750 | 1.720 | 50.0 | 0.667 | 1.750 | 33.3 | 16.7 | -0.833 | 1.720 | 16.7 | 0.0 |
| Income |  |  |  |  |  |  |  |  |  |  |  |
| < \$20,000 | 0.672 | 1.570 | 47.8 | 0.380 | 1.650 | 37.7 | 26.8 | -0.964 | 1.640 | 17.4 | 6.9 |
| \$20,000-\$40,000 | 0.480 | 1.700 | 46.6 | 0.215 | 1.730 | 37.1 | 25.0 | -0.745 | 1.790 | 27.8 | 10.8 |
| \$40,000-\$60,000 | 0.592 | 1.630 | 49.4 | 0.220 | 1.670 | 36.9 | 25.4 | -0.930 | 1.750 | 20.5 | 8.9 |
| \$60,000-\$80,000 | 0.629 | 1.620 | 49.5 | 0.376 | 1.640 | 38.0 | 27.4 | -0.883 | 1.710 | 20.9 | 10.5 |
| \$80,000-\$100,000 | 0.741 | 1.520 | 50.0 | 0.488 | 1.530 | 41.3 | 27.2 | -0.994 | 1.640 | 18.9 | 6.0 |
| > \$100,000 | 0.608 | 1.620 | 47.2 | 0.302 | 1.680 | 37.5 | 25.7 | -0.914 | 1.700 | 21.0 | 7.4 |
| Prefer not to answer | 0.861 | 1.940 | 47.2 | 0.556 | 2.080 | 38.9 | 36.1 | -1.170 | 1.930 | 19.4 | 2.8 |
| No response | -0.250 | 0.866 | 25.0 | -0.500 | 1.000 | 0.0 | 0.0 | 0.000 | 0.816 | 25.0 | 0.0 |
|  | mean | SD | % | mean | SD | % | % | mean | SD | % | % |
| Political Ideology |  |  |  |  |  |  |  |  |  |  |  |
| Very liberal | 0.888 | 1.740 | 54.3 | 0.590 | 1.780 | 44.1 | 31.1 | -1.190 | 1.830 | 19.8 | 6.1 |
| Liberal | 0.753 | 1.650 | 51.6 | 0.491 | 1.680 | 42.3 | 29.8 | -1.010 | 1.740 | 20.2 | 8.2 |
| Moderate | 0.557 | 1.570 | 47.5 | 0.247 | 1.600 | 36.2 | 25.4 | -0.867 | 1.670 | 21.1 | 8.1 |
| Conservative | 0.380 | 1.600 | 43.8 | 0.058 | 1.650 | 33.1 | 21.4 | -0.703 | 1.700 | 25.0 | 11.2 |
| Very conservative | 0.307 | 1.520 | 39.0 | 0.026 | 1.570 | 27.7 | 18.6 | -0.589 | 1.500 | 24.2 | 9.5 |
| Prefer not to answer | 0.684 | 1.680 | 57.9 | 0.263 | 1.560 | 31.6 | 21.1 | -1.110 | 1.940 | 21.1 | 15.8 |
| No response | 0.625 | 0.750 | 50.0 | 0.250 | 0.957 | 50.0 | 50.0 | -1.000 | 0.816 | 0.0 | 0.0 |
| Political Ideology (Social) |  |  |  |  |  |  |  |  |  |  |  |
| Very liberal | 0.927 | 1.720 | 55.7 | 0.628 | 1.760 | 46.3 | 33.3 | -1.230 | 1.810 | 19.1 | 5.5 |
| Liberal | 0.714 | 1.610 | 51.2 | 0.445 | 1.640 | 41.1 | 28.5 | -0.983 | 1.710 | 20.9 | 8.2 |
| Moderate | 0.498 | 1.600 | 45.2 | 0.205 | 1.660 | 35.2 | 25.0 | -0.791 | 1.680 | 22.1 | 9.4 |
| Conservative | 0.321 | 1.590 | 42.5 | -0.016 | 1.630 | 30.6 | 19.8 | -0.658 | 1.710 | 25.1 | 12.1 |
| Very conservative | 0.362 | 1.500 | 40.6 | 0.059 | 1.550 | 28.9 | 18.8 | -0.665 | 1.590 | 22.6 | 8.0 |
| Prefer not to answer | 0.528 | 1.540 | 55.6 | 0.222 | 1.560 | 33.3 | 11.1 | -0.833 | 1.650 | 16.7 | 11.1 |
| No response | -1.000 | NA | 0.0 | -2.000 | NA | 0.0 | 0.0 | 0.000 | NA | 0.0 | 0.0 |
| Political Ideology (Economic) |  |  |  |  |  |  |  |  |  |  |  |
| Very liberal | 0.795 | 1.760 | 49.4 | 0.514 | 1.770 | 40.5 | 28.6 | -1.080 | 1.870 | 19.9 | 6.7 |
| Liberal | 0.800 | 1.630 | 53.8 | 0.512 | 1.670 | 43.7 | 31.5 | -1.090 | 1.730 | 18.9 | 7.8 |
| Moderate | 0.594 | 1.600 | 48.2 | 0.307 | 1.650 | 38.0 | 25.5 | -0.882 | 1.670 | 21.4 | 8.4 |
| Conservative | 0.401 | 1.580 | 44.2 | 0.076 | 1.620 | 33.5 | 22.4 | -0.726 | 1.710 | 25.5 | 10.4 |
| Very conservative | 0.435 | 1.600 | 42.9 | 0.165 | 1.650 | 30.7 | 21.7 | -0.705 | 1.660 | 22.7 | 9.6 |
| Prefer not to answer | 0.783 | 1.540 | 65.2 | 0.435 | 1.530 | 39.1 | 21.7 | -1.130 | 1.660 | 13.0 | 8.7 |
| No response | -1.000 | 0.000 | 0.0 | -1.500 | 0.707 | 0.0 | 0.0 | 0.500 | 0.707 | 50.0 | 0.0 |
| Political Party |  |  |  |  |  |  |  |  |  |  |  |
| Strong Democrat | 0.869 | 1.710 | 54.6 | 0.582 | 1.720 | 43.9 | 28.7 | -1.160 | 1.820 | 19.6 | 7.6 |
| Democrat | 0.701 | 1.630 | 50.7 | 0.411 | 1.690 | 39.7 | 29.9 | -0.990 | 1.700 | 19.9 | 6.7 |
| Independent (but lean Democrat) | 0.755 | 1.620 | 51.9 | 0.470 | 1.640 | 42.0 | 29.6 | -1.040 | 1.730 | 21.0 | 8.6 |
| Independent | 0.468 | 1.590 | 43.7 | 0.173 | 1.630 | 34.0 | 23.3 | -0.762 | 1.670 | 22.1 | 9.2 |
| Independent (but lean Republican) | 0.437 | 1.720 | 42.4 | 0.144 | 1.730 | 33.9 | 24.7 | -0.731 | 1.830 | 28.8 | 14.8 |
| Republican | 0.387 | 1.550 | 44.8 | 0.076 | 1.610 | 33.4 | 20.9 | -0.699 | 1.640 | 22.5 | 8.8 |
| Strong Republican | 0.432 | 1.500 | 44.0 | 0.130 | 1.570 | 32.6 | 20.7 | -0.734 | 1.580 | 21.7 | 7.6 |
| Prefer not to answer | 0.615 | 1.580 | 56.4 | 0.282 | 1.490 | 41.0 | 23.1 | -0.949 | 1.790 | 20.5 | 10.3 |
| No response | -1.000 | NA | 0.0 | -2.000 | NA | 0.0 | 0.0 | 0.000 | NA | 0.0 | 0.0 |
|  | mean | SD | % | mean | SD | % | % | mean | SD | % | % |
| Religion |  |  |  |  |  |  |  |  |  |  |  |
| Christian - Protestant | 0.515 | 1.620 | 45.9 | 0.212 | 1.680 | 34.9 | 24.3 | -0.818 | 1.700 | 22.5 | 10.0 |
| Christian - Catholic | 0.483 | 1.510 | 46.7 | 0.176 | 1.550 | 34.4 | 21.6 | -0.790 | 1.610 | 20.7 | 6.4 |
| Christian - Other | 0.589 | 1.650 | 48.3 | 0.298 | 1.690 | 37.3 | 25.4 | -0.881 | 1.740 | 22.9 | 9.7 |
| Jewish | 0.868 | 1.720 | 54.7 | 0.453 | 1.840 | 43.4 | 32.1 | -1.280 | 1.770 | 13.2 | 7.6 |
| Muslim | 0.357 | 1.700 | 45.7 | -0.057 | 1.800 | 28.6 | 20.0 | -0.771 | 1.780 | 31.4 | 17.1 |
| Buddhist | 0.840 | 1.690 | 54.0 | 0.520 | 1.570 | 48.0 | 32.0 | -1.160 | 1.940 | 24.0 | 14.0 |
| Hindu | -0.129 | 1.550 | 38.7 | -0.452 | 1.570 | 29.0 | 16.1 | -0.194 | 1.620 | 35.5 | 19.4 |
| Non-religious | 0.704 | 1.650 | 49.9 | 0.435 | 1.680 | 40.7 | 28.5 | -0.973 | 1.750 | 21.1 | 8.0 |
| Other | 0.673 | 1.780 | 49.0 | 0.337 | 1.810 | 40.4 | 31.7 | -1.010 | 1.880 | 22.1 | 8.7 |
| Prefer not to answer | 1.090 | 1.570 | 58.8 | 0.794 | 1.650 | 41.2 | 38.2 | -1.380 | 1.600 | 11.8 | 0.0 |
| No response | 1.250 | 1.770 | 50.0 | 1.000 | 1.410 | 50.0 | 50.0 | -1.500 | 2.120 | 0.0 | 0.0 |
| STEM degree |  |  |  |  |  |  |  |  |  |  |  |
| No | 0.587 | 1.620 | 47.9 | 0.289 | 1.650 | 37.2 | 25.6 | -0.885 | 1.720 | 21.3 | 8.4 |
| Yes | 0.680 | 1.680 | 49.8 | 0.397 | 1.740 | 40.3 | 28.5 | -0.963 | 1.750 | 22.9 | 10.0 |
| Prefer not to answer | 0.400 | 1.510 | 40.0 | 0.200 | 1.510 | 30.0 | 15.0 | -0.600 | 1.570 | 25.0 | 0.0 |
| No response | 0.250 | 1.060 | 50.0 | -0.500 | 0.707 | 0.0 | 0.0 | -1.000 | 1.410 | 0.0 | 0.0 |

**Table S10.** Correlations between clinician characteristics and sentiments about experiments.

|  | Size of A/B effect |  | A/B effect |  | Size of experiment aversion |  | Experiment aversion |  | Experiment rejection |  | Size of experiment appreciation |  | Experiment appreciation |  | Experiment endorsement |  |
| --- | --- | --- | --- | --- | --- | --- | --- | --- | --- | --- | --- | --- | --- | --- | --- | --- |
|  | *r* | *p* | *r* | *p* | *r* | *p* | *r* | *p* | *r* | *p* | *r* | *p* | *r* | *p* | *r* | *p* |
| Sex<br>(1 = male, 2 = female) | 0.016 | 0.453 | 0.016 | 0.457 | 0.000 | 0.991 | -0.011 | 0.619 | -0.021 | 0.326 | -0.030 | 0.165 | -0.026 | 0.226 | -0.032 | 0.134 |
| Number of research methods/statistics training units | -0.005 | 0.812 | 0.000 | 0.992 | 0.000 | 0.999 | 0.016 | 0.471 | 0.017 | 0.428 | 0.010 | 0.659 | 0.019 | 0.382 | 0.010 | 0.643 |
| Comfort with research methods/statistics | -0.036 | 0.100 | -0.018 | 0.410 | -0.039 | 0.071 | -0.021 | 0.335 | -0.016 | 0.446 | 0.030 | 0.165 | 0.070 | 0.001 | 0.045 | 0.035 |
| Number of research methods/statistics activities | -0.019 | 0.375 | -0.022 | 0.301 | -0.006 | 0.796 | 0.006 | 0.778 | 0.020 | 0.360 | 0.031 | 0.157 | 0.041 | 0.056 | 0.023 | 0.279 |
| Currently involved in research | -0.002 | 0.912 | -0.012 | 0.570 | -0.009 | 0.691 | -0.016 | 0.470 | -0.022 | 0.309 | -0.004 | 0.870 | -0.024 | 0.267 | 0.009 | 0.693 |
| Position<br>(0 = non-prescriber, 1 = prescriber) | 0.033 | 0.121 | 0.029 | 0.176 | 0.040 | 0.061 | 0.042 | 0.050 | 0.052 | 0.016 | -0.025 | 0.250 | -0.020 | 0.347 | -0.021 | 0.338 |
| Years in medicine | 0.016 | 0.452 | -0.004 | 0.865 | 0.011 | 0.599 | -0.007 | 0.734 | 0.006 | 0.792 | -0.020 | 0.362 | 0.029 | 0.185 | -0.003 | 0.879 |

**Table S11.** Means and percentages of sentiments about experiments by demographic variable in clincian sample.

|  | Size of A/B effect |  | A/B effect | Size of experiment aversion |  | Experiment aversion | Experiment rejection | Size of experiment appreciation |  | Experiment appreciation | Experiment endorsement |
| --- | --- | --- | --- | --- | --- | --- | --- | --- | --- | --- | --- |
|  | mean | SD | % | mean | SD | % | % | mean | SD | % | % |
| Sex |  |  |  |  |  |  |  |  |  |  |  |
| Male | 0.456 | 1.800 | 43.9 | 0.270 | 1.800 | 38.5 | 28.2 | -0.642 | 1.890 | 26.5 | 17.2 |
| Female | 0.529 | 1.750 | 45.9 | 0.271 | 1.750 | 37.2 | 25.8 | -0.786 | 1.890 | 23.6 | 14.2 |
| Other | 0.000 | 1.870 | 40.0 | 0.000 | 1.870 | 40.0 | 20.0 | 0.000 | 1.870 | 20.0 | 20.0 |
| Source of research methods/statistics training |  |  |  |  |  |  |  |  |  |  |  |
| Undergraduate coursework | 0.483 | 1.755 | 44.2 | 0.258 | 1.753 | 37.7 | 26.5 | -0.707 | 1.870 | 25.0 | 14.1 |
| Professional school instruction | 0.571 | 1.767 | 46.0 | 0.314 | 1.756 | 38.2 | 27.1 | -0.828 | 1.916 | 22.8 | 14.7 |
| Postgraduate coursework | 0.624 | 1.818 | 49.4 | 0.402 | 1.809 | 41.5 | 29.4 | -0.847 | 1.936 | 24.5 | 14.5 |
| CME/CEU courses | 0.463 | 1.788 | 47.1 | 0.217 | 1.767 | 38.6 | 26.6 | -0.708 | 1.925 | 25.7 | 16.7 |
| Self-instruction via peer-reviewed literature | 0.333 | 1.820 | 41.2 | 0.097 | 1.798 | 32.9 | 23.2 | -0.569 | 1.949 | 27.3 | 16.6 |
| Other | 0.722 | 1.902 | 46.7 | 0.478 | 1.915 | 41.1 | 32.2 | -0.967 | 1.986 | 22.2 | 14.4 |
| Comfort with research methods/statistics |  |  |  |  |  |  |  |  |  |  |  |
| Not at all | 0.682 | 1.760 | 45.8 | 0.432 | 1.780 | 37.7 | 26.3 | -0.932 | 1.870 | 18.2 | 12.7 |
| Somewhat | 0.516 | 1.710 | 45.7 | 0.282 | 1.690 | 37.8 | 26.8 | -0.750 | 1.840 | 22.5 | 14.0 |
| Moderately | 0.482 | 1.770 | 46.5 | 0.237 | 1.770 | 38.3 | 26.6 | -0.727 | 1.880 | 26.8 | 15.1 |
| Very | 0.491 | 1.910 | 43.9 | 0.203 | 1.900 | 34.0 | 23.1 | -0.778 | 2.070 | 29.2 | 17.9 |
| Extremely | 0.105 | 2.020 | 31.6 | -0.079 | 2.050 | 28.9 | 23.7 | -0.289 | 2.100 | 26.3 | 23.7 |
| Research methods/statistics activities |  |  |  |  |  |  |  |  |  |  |  |
| Read results of RCT in peer-reviewed journal article | 0.521 | 1.772 | 45.5 | 0.284 | 1.762 | 38.0 | 27.2 | -0.758 | 1.898 | 24.7 | 15.0 |
| Changed typical prescription/recommendation after personally reading results of RCT in peer-reviewed journal article | 0.430 | 1.813 | 43.3 | 0.217 | 1.814 | 36.8 | 26.3 | -0.643 | 1.921 | 26.6 | 16.7 |
| Published scientific paper in peer-reviewed journal | 0.530 | 1.692 | 43.3 | 0.339 | 1.681 | 38.2 | 29.9 | -0.720 | 1.802 | 22.8 | 13.4 |
| Conducted or worked on a team conducting an RCT | 0.371 | 1.745 | 42.9 | 0.114 | 1.725 | 35.1 | 20.9 | -0.628 | 1.902 | 25.8 | 16.3 |
| Took a course/class in statistics, biostatistics, research methods | 0.505 | 1.775 | 45.0 | 0.277 | 1.770 | 37.8 | 27.3 | -0.732 | 1.892 | 25.4 | 15.2 |
| Analyzed data for statistical significance outside of course requirement | 0.470 | 1.781 | 43.7 | 0.251 | 1.766 | 36.7 | 26.2 | -0.690 | 1.912 | 26.2 | 15.4 |
| Used statistical software | 0.588 | 1.803 | 49.3 | 0.389 | 1.795 | 42.5 | 31.7 | -0.787 | 1.915 | 26.7 | 14.9 |

|  | Size of<br>A/B<br>effect |  | A/B effect | Size of<br>experiment<br>aversion |  | Experiment<br>aversion | Experiment<br>rejection | Size of<br>experiment<br>appreciation |  | Experiment<br>appreciation | Experiment<br>endorsement |
| --- | --- | --- | --- | --- | --- | --- | --- | --- | --- | --- | --- |
|  | mean | SD | % | mean | SD | % | % | mean | SD | % | % |
| Currently involved in research |  |  |  |  |  |  |  |  |  |  |  |
| Yes | 0.526 | 1.740 | 47.4 | 0.316 | 1.720 | 39.7 | 29.2 | -0.737 | 1.860 | 27.3 | 13.9 |
| No | 0.512 | 1.760 | 45.3 | 0.265 | 1.760 | 37.2 | 25.9 | -0.759 | 1.890 | 23.8 | 14.9 |
| Position |  |  |  |  |  |  |  |  |  |  |  |
| Doctor | 0.556 | 1.730 | 45.5 | 0.374 | 1.720 | 39.9 | 28.7 | -0.738 | 1.840 | 23.1 | 13.7 |
| Physician Assistant | 0.757 | 1.780 | 53.0 | 0.508 | 1.780 | 44.3 | 34.4 | -1.010 | 1.890 | 21.9 | 13.1 |
| Nurse Practitioner | 0.500 | 1.910 | 45.9 | 0.184 | 1.970 | 36.7 | 25.5 | -0.816 | 2.030 | 23.5 | 14.3 |
| Nurse (RN) | 0.436 | 1.720 | 43.8 | 0.181 | 1.720 | 35.2 | 23.9 | -0.690 | 1.850 | 25.3 | 15.1 |
| Nurse (LPN) | 0.410 | 1.790 | 42.1 | 0.150 | 1.760 | 33.5 | 22.6 | -0.669 | 1.960 | 24.8 | 17.3 |
| Nurse (Other) | 1.180 | 1.910 | 65.0 | 0.800 | 1.910 | 55.0 | 35.0 | -1.550 | 2.060 | 10.0 | 10.0 |
| Genetic Counselor | --- | --- | --- | --- | --- | --- | --- | --- | --- | --- | --- |
| Non-prescribing clinician or staff without clinical<br>credential | --- | --- | --- | --- | --- | --- | --- | --- | --- | --- | --- |
| Medical student | 1.170 | 1.770 | 65.2 | 0.935 | 1.790 | 56.5 | 45.7 | -1.410 | 1.830 | 15.2 | 8.7 |
| Faculty or Professor | 1.120 | 2.050 | 62.5 | 0.875 | 2.030 | 50.0 | 37.5 | -1.380 | 2.200 | 25.0 | 12.5 |
| Other | 0.727 | 2.000 | 45.5 | 0.618 | 1.980 | 41.8 | 32.7 | -0.836 | 2.060 | 25.5 | 16.4 |
| Years in medical field |  |  |  |  |  |  |  |  |  |  |  |
| < 1 year | 0.582 | 1.540 | 47.5 | 0.377 | 1.540 | 39.3 | 32.8 | -0.787 | 1.660 | 24.6 | 8.2 |
| 1-2 years | 0.560 | 1.720 | 48.4 | 0.333 | 1.710 | 41.3 | 29.4 | -0.786 | 1.840 | 23.8 | 14.3 |
| 3-5 years | 0.392 | 1.570 | 44.8 | 0.140 | 1.570 | 36.0 | 21.3 | -0.643 | 1.690 | 23.4 | 13.6 |
| 6-10 years | 0.423 | 1.730 | 43.3 | 0.205 | 1.760 | 36.5 | 24.6 | -0.641 | 1.830 | 26.4 | 15.1 |
| > 10 years | 0.555 | 1.820 | 45.9 | 0.303 | 1.810 | 37.5 | 27.1 | -0.807 | 1.950 | 23.7 | 15.3 |

In the lay sample, women show larger AB and experiment aversion effects (e.g., larger difference between mean intervention rating/lowest-rated intervention rating and AB test rating; *r* = .067–.068, *p* < .001) and a smaller experiment appreciation effect (e.g., smaller difference between AB test and highest-rated intervention rating; *r* = –.064, *p* < .001). Lay participants who are more conservative (in general and with respect to social and economic issues) or more likely to be strong Republicans show lower levels of an AB effect and experiment aversion (i.e., smaller difference between mean intervention rating/lowest-rated intervention rating and AB test rating; all *r*s < –.094, *p*s < .0001). These participants also show significantly more experiment appreciation, though the strength of the association is weaker (*r*s = .037–.046, *p* < .0001). Finally, we find that people who are non-religious show a larger degree of experiment aversion (*r* = .061, *p* < .001; they also show a larger AB effect, *r* = .051, but *p* = .007 which is greater than *p* < .005, the standard proposed in Benjamin et al. (2018)^17^ for exploratory analyses without a priori hypotheses). For all other variables, we find no significant associations between the individual difference measures and experiment sentiments (all *r*s < |.051|, all *p*s > .005).

In the clinician sample, the strongest association was between self-reported comfort with research methods and statistics and experiment aversion—clinicians who report being more comfortable with research methods and statistics are more likely to appreciate the A/B test (*r* = .070, *p* = .001).

## Footnotes

1 The Best Vaccine vignette was combined with another study that required a sample size much larger than the sample sizes in our previous vignette studies to have adequate statistical power.

2 For the clinician study of the Intubation Safety Checklist, Best Corticosteroid Drug, and Masking Rules vignettes, clinicians were randomly assigned to one of these three scenarios and then completed the remaining two scenarios in random order. For consistency with the rest of this project and with our previous survey experiment with clinicians regarding the A/B effect (Meyer et al., 2019, Study 6), and in order to make the results from clinician samples comparable to those with lay samples (in which each participant only ever saw one scenario), we analyze data from this study as a between-subjects design where we only consider the first scenario that every participant completed. See the section “Order Effect in Clinician Study” elsewhere in this appendix for further analyses.

3 The clinician version of the Best Vaccine vignette was combined with another study being conducted by a subset of researchers on this team. The materials for Best Vaccine were presented after the survey materials from the other study. Data from the other study are unrelated to the research questions tested here and will be reported separately.

4 In all vignettes, the protagonist (e.g., the hospital director or Dr. Jones) was male for ease of comparison to our previous work using these vignettes. Future work should examine the impact of the characteristics of the decision-maker on evaluations of their decisions regarding policy imposition and conducting RCTs.

